# Immunogenicity and safety of SARS-CoV-2 recombinant protein nanoparticle vaccine GBP510 adjuvanted with AS03: randomised, active-controlled, observer-blinded, phase 3 trial

**DOI:** 10.1101/2023.01.31.23284895

**Authors:** Joon Young Song, Won Suk Choi, Jung Yeon Heo, Eun Jin Kim, Jin Soo Lee, Dong Sik Jung, Shin-Woo Kim, Kyung-Hwa Park, Joong Sik Eom, Su Jin Jeong, Jacob Lee, Ki Tae Kwon, Hee Jung Choi, Jang Wook Sohn, Young Keun Kim, Byung Wook Yoo, In-Jin Jang, Maria R. Capeding, François Roman, Thomas Breuer, Piotr Wysocki, Lauren Carter, Sushant Sahastrabuddhe, Manki Song, Naveena D’Cor, Hun Kim, Ji Hwa Ryu, Su Jeen Lee, Yong Wook Park, Hee Jin Cheong

**Affiliations:** Division of Infectious Diseases, Department of Internal Medicine, Korea University Guro Hospital, Korea University College of Medicine, Seoul, Republic of Korea (JY Song, HJ Cheong); Division of Infectious Diseases, Department of Internal Medicine, Korea University Ansan Hospital, Korea University College of Medicine, Ansan, Republic of Korea (WS Choi); Department of Infectious Diseases, Ajou University School of Medicine, Suwon, Republic of Korea (JY Heo, EJ Kim); Division of Infectious Diseases, Department of Internal Medicine, Inha University College of Medicine, Incheon, Republic of Korea (JS Lee); Division of Infectious Diseases, Department of Internal Medicine, Dong-A University College of Medicine, Busan, Republic of Korea (DS Jung); Division of Infectious Diseases, Department of Internal Medicine, Kyungpook National University Hospital, School of Medicine, Kyungpook National University, Daegu, Republic of Korea (SW Kim); Division of Infectious Diseases, Department of Internal Medicine, Chonnam National University Medical School, Gwangju, Republic of Korea (KH Park); Division of Infectious Diseases, Department of Internal Medicine, Gil Medical Center, Gachon University College of Medicine, Incheon, Republic of Korea (JS Eom); Division of Infectious Diseases, Department of Internal Medicine, Severance Hospital, Yonsei University College of Medicine, Seoul, Republic of Korea (SJ Jeong); Division of Infectious Diseases, Department of Internal Medicine, Hallym University College of Medicine, Chuncheon, Republic of Korea (J Lee); Division of Infectious Diseases, Department of Internal Medicine, Kyungpook National University Chilgok Hospital, School of Medicine, Kyungpook National University, Daegu, Republic of Korea (KT Kwon); Division of Infectious Diseases, Department of Internal Medicine, Ewha Womans University Mokdong Hospital, Seoul, Republic of Korea (HJ Choi); Division of Infectious Diseases, Department of Internal Medicine, Korea University Anam Hospital, Korea University College of Medicine, Seoul, Republic of Korea (JW Sohn); Division of Infectious Diseases, Department of Internal Medicine, Yonsei University Wonju Severance Christian; Hospital, Yonsei University Wonju College of Medicine, Wonju, Republic of Korea (YK Kim); Department of Family Medicine, Soon Chun Hyang University Hospital, Seoul, Republic of Korea (BW Yoo); Department of Clinical Pharmacology and Therapeutics, College of Medicine, Seoul National University Hospital, Seoul, Republic of Korea (IJ Jang); Tropical Disease Foundation – San Francisco Multi-purpose Bldg, Laguna 4000, Philippines (M. R. Capeding); GlaxoSmithKline Vaccines, Wavre, Belgium (F Roman, T Breuer); GlaxoSmithKline Vaccines, Warsaw, Poland (P Wysocki); Department of Biochemistry, University of Washington, WA, U.S. (L Carter); Institute for Protein Design, University of Washington, WA, U.S. (L Carter); International Vaccine Institute, Seoul, Republic of Korea (S Sahastrabudhe, M Song, N D’Cor); Department of R&D, SK Bioscience, Seongnam, Republic of Korea (H Kim, JH Ryu, SJ Lee, YW Park)

**Author notes:** Corresponding author: Hee Jin Cheong, MD, PhD, Division of Infectious Diseases, Department of Internal Medicine, Korea University Guro Hospital, Korea University College of Medicine, Gurodong-ro 148, Guro-gu, Seoul 08308, Republic of Korea.

## Abstract

**Background:** GBP510 vaccine contains self-assembling, recombinant nanoparticles displaying SARS-CoV-2 spike receptor-binding domains. We report interim phase 3 immunogenicity results for GBP510 adjuvanted with AS03 (GBP510/AS03) compared with ChAdOx1-S (Vaxzevria, AstraZeneca) up to 2 weeks after the second dose, and safety data up to a median of 2.5 months follow-up.

**Methods:** Randomised, active-controlled, observer-blinded, multinational study: Cohort 1 (no history of SARS-CoV-2 infection/COVID-19 vaccination, n=1956) randomised 2:1 to receive two doses of GBP510/AS03 or ChAdOx1-S (immunogenicity and safety); Cohort 2 (regardless of baseline serostatus; n=2080) randomised 5:1 (safety). Primary objectives: demonstrate superiority in geometric mean titre (GMT) and non-inferiority in seroconversion rate (SCR; ≥4-fold rise from baseline) of GBP510/AS03 versus ChAdOx1-S for neutralising antibodies against the ancestral strain by live-virus neutralisation assay. Secondary objectives included assessment of safety and reactogenicity.

**Findings:** At 2 weeks after the second vaccination, the GMT ratio (GBP510/AS03 / ChAdOx1-S) was 2.93 (95% CI 2.63–3.27), demonstrating superiority (95% CI lower limit >1). The between-group SCR difference of 10.76% (95% CI 7.68–14.32) satisfied the non-inferiority criterion (95% CI lower limit > −5%).The proportion of subjects with adverse events (AEs) after any vaccination was higher with GBP510/AS03 versus ChAdOx1-S for solicited local AEs (56.69% vs 49.20%), but was similar for solicited systemic AEs (51.21% vs 53.51%) and unsolicited AEs (13.27% vs 14.56%). No safety concerns were identified during follow-up for a median 2.5-months after the second vaccination.

**Interpretation:** GBP510/AS03 met the superiority criterion for neutralising antibodies and non-inferiority criterion for SCR compared with ChAdOx1-S, and showed a clinically acceptable safety profile.

**Funding:** This work was supported, in whole or in part, by funding from CEPI and the Bill & Melinda Gates Foundation Investments INV-010680 and INV-006462. The Bill & Melinda Gates Foundation supported this project for the generation of IND-enabling data and CEPI supported this clinical study.

**RESEARCH IN CONTEXT:** *Evidence before this study:* Immunobridging has been proposed as an approach for assessing new COVID-19 vaccines by comparing the immunogenicity of candidate vaccines with an active comparator with demonstrated clinical efficacy. We searched PubMed up to 26 October 2022 for immunobridging clinical trials comparing a candidate vaccine with an approved vaccine, using the terms “immunobridging”, “SARS-CoV-2”, “COVID-19”, and “vaccine”. A phase 2/3 study showed that the ChAdOx1 vaccine, manufactured by the Serum Institute of India after technology transfer from Oxford University/AstraZeneca, had a non-inferior immune response compared to the original ChAdOx1 (AZD1222) vaccine. A post hoc analysis of phase 2 data found that MVC-COV1901 vaccine (a protein subunit vaccine developed by Medigen Vaccine Biologics Corporation, Taiwan) was non-inferior to ChAdOx1 (AZD1222) with respect to neutralising antibody titres. A phase 3 study found that VLA2001 (an adjuvanted, inactivated whole-virus vaccine developed by Valneva, Austria) was superior to ChAdOx1 with respect to neutralising antibody titres and non-inferior with respect to seroconversion rates.

*Added value of this study:* This is the first study comparing the immunogenicity of recombinant SARS-CoV-2 protein nanoparticle vaccine GBP510 adjuvanted with AS03 versus ChAdOx1-S. Interim analysis found that two-dose vaccination with GBP510/AS03 induced stronger neutralising antibody immune responses compared with ChAdOx1-S against the ancestral D614G strain at 2 weeks after the second dose. GBP510/AS03 had an acceptable safety profile during a median 2.5 months of follow-up.

*Implications of the available evidence:* This interim analysis suggests that GBP510/AS03 induces strong neutralising antibody responses against SARS-CoV-2 ancestral strain and has an acceptable safety profile.

## INTRODUCTION

Multiple vaccines against severe acute respiratory syndrome coronavirus 2 (SARS-CoV-2), based on different underlying technologies, have been approved in different countries.^1^

GBP510 is a recombinant protein vaccine consisting of self-assembling, two-component nanoparticles displaying SARS-CoV-2 spike receptor-binding domains (RBDs). It is adjuvanted with AS03, which contains α-tocopherol and squalene^2^ and enhances the immune response to the vaccine antigen.^3^ The vaccine can be stored at regular refrigerator temperatures (2–8 ℃) which makes it suitable for rollout in parts of the world where the requirement for ultra-cold chain handling can be challenging. A phase 1/2 study showed that GBP510 adjuvanted with AS03 (hereafter GBP510/AS03) was highly immunogenic and well tolerated in healthy adults aged 19–85 years.^3^

Thresholds for immune correlates of protection based on antibody levels or functional activity have not yet been established.^4^ However, in 2021, the International Coalition of Medicines Regulatory Authorities determined that immunobridging studies were acceptable as part of the strategy for assessing new COVID-19 vaccines,^5^ a position which has since been adopted by regulatory bodies, including members of the Access Consortium (national regulatory authorities of Australia, Canada, Singapore, Switzerland and the UK).^6^

The aim of the current study was to assess the immunogenicity and safety of GBP510/AS03 for the prevention of COVID-19, based on the assumption that neutralising antibody titres would predict efficacy against the parental D614G strain. The primary objective was to demonstrate that the immune response induced by two doses of GBP510/AS03 administered at a 1-month interval in seronegative adults was superior (based on geometric mean titre [GMT] of neutralising antibodies) and non-inferior (based on seroconversion rate [SCR]) to the immune response induced by two doses of the ChAdOx1-S vaccine (Vaxzevria, AstraZeneca)^7^ against the ancestral strain using live virus neutralising assays. The secondary objective was to assess the safety profile of GBP510/AS03 regardless of baseline serostatus.

Follow-up of participants until 12 months after their second vaccination is ongoing. The interim analysis reported here encompasses immunogenicity data up to 2 weeks and safety data up to a median of 2.5 months follow-up after the second dose (data cut-off 31 March 2022).

## METHODS

### Study design and participants

This was a randomised, active-controlled, observer-blinded, parallel group, phase 3 study, conducted at 38 sites across six countries (South Korea, Philippines, Thailand, Vietnam, Ukraine and New Zealand).

Healthy or medically stable adults aged ≥18 years were enrolled into one of two cohorts (Figure S1). Cohort 1 (Immunogenicity cohort) included subjects with no history of SARS-CoV-2 infection or COVID-19 vaccination (confirmed by a rapid antibody screening test). Cohort 2 (Safety cohort) enrolled subjects irrespective of their serostatus at screening. Subjects were excluded from either cohort if they met any of the following criteria: clinically significant respiratory symptoms, febrile illness, or acute illness within 72h; previous SARS or middle east respiratory syndrome (MERS) infection, or SARS/MERS vaccination; receipt of medications/vaccinations aimed at preventing COVID-19; immunocompromised conditions; autoimmune diseases; bleeding disorders, thrombocytopenia, thrombosis or capillary leak syndrome; malignancy within 1 year; hypersensitivity to any vaccines; receipt of any other vaccine between 4 weeks(2 weeks for influenza vaccines) before first and 28 days after last study vaccination; receipt of immunoglobulins, blood products or systemic corticosteroids within 12 weeks; meeting any restriction for ChAdOx1-S; significant unstable chronic illness; pregnancy or breast feeding.

The study was designed by SK Bioscience with support from GlaxoSmithKline (GSK), the International Vaccine Institute and the Coalition for Epidemic Preparedness Innovations (CEPI). The study was performed in accordance with the Declaration of Helsinki and the International Council for Harmonization Good Clinical Practice guidelines. The protocol (supplementary material) was approved by the Institutional Review Board for each participating health facility, and written informed consent was obtained from all participants. The reporting of this study complies with the Consolidated Standards of Reporting Trials (CONSORT) 2010 Statement. This study is registered on ClinicalTrials.gov (NCT05007951).

### Randomisation and masking

For Cohort 1 sites, participants were randomised in a 2:1 ratio to receive GBP510/AS03 or ChAdOx1-S. For Cohort 2 sites, participants were randomised in a 5:1 ratio to GBP510/AS03 or ChAdOx1-S. Centralised interactive response technology was used to randomly allocate participants to treatment according to pre-generated block randomisation schedules stratified by age (18–64 or ≥65 years) and trial site. Except for pharmacy staff and vaccinators, all other study and laboratory personnel, and participants were blinded to treatment assignment.

### Procedures

The study vaccine GBP510, which contains self-assembling, two-component nanoparticles (RBD-16GS-I53-50) that display SARS-CoV-2 spike (S) protein RBDs, was developed by the Institute for Protein Design at the University of Washington and SK Bioscience. The AS03 adjuvant (an α-tocopherol-containing oil-in-water emulsion) was developed by GlaxoSmithKline.^2^ The control vaccine ChAdOx1-S was Chimpanzee Adenovirus encoding the SARS-CoV-2 spike glycoprotein (Vaxzevria, AstraZeneca).^8^

Each participant received two intramuscular injections (0.5 mL volume) of vaccine into the deltoid muscle at a 28-day interval. Each dose of the GBP510/AS03 vaccine contained RBD 25 μg (0.25 mL) adjuvanted with AS03 (0.25 mL). Each dose of ChAdOx1-S contained no less than 2.5×10^8^ infectious units/0.5mL.

Safety evaluations were performed for all participants who received at least one dose of study intervention. Immunogenicity assessments were performed in Cohort 1 only.

All participants were observed for at least 30 minutes after each vaccination for immediate adverse events. Participants used diary cards to record solicited local adverse events (AEs) (redness, swelling, and pain at injection site) and solicited systemic AEs (fever, nausea/vomiting, diarrhoea, fatigue, myalgia, arthralgia, chills) for 7 days after each vaccination, and unsolicited AEs within 28 days after each vaccination. Serious AEs (SAEs), medically-attended AEs (MAAEs), AEs leading to study withdrawal, and AEs of special interest (AESIs; Tables S1 and S2) were recorded throughout the entire study period. AE severity was based on the US Food and Drug Administration (FDA) toxicity grading scale.^9^ All solicited local and systemic AEs were considered related to the study intervention; the causal relationship for unsolicited AE was assessed by the investigators. Safety data were reviewed during the study by an independent safety monitoring board.

Blood samples for assessment of IgG antibody response and neutralisation assays were planned to be obtained from Cohort 1 participants at baseline, 4 weeks after the first vaccination, and at 2 weeks, 4 weeks, 6 months, and 12 months after the second vaccination. Blood samples for cell-mediated immunity assessments were collected at baseline and 2 weeks, 6 months, and 12 months after the second vaccination from a subset of approximately 10% of Cohort 1, selected pragmatically in advance to retain the randomisation ratio between study groups.

The neutralising antibody response to SARS-CoV-2 was measured using a focus reduction neutralisation test (FRNT) which assesses neutralising antibody titres in serum induced by viral infection/vaccination with live virus. The IgG antibody response to SARS-CoV-2 RBD was measured using enzyme-linked immunosorbent assay (ELISA). Cell-mediated immune responses were assessed using FluoroSpot assays to measure cytokine secretion in cells induced by external antigens such as viruses or vaccines, and intracellular cytokine staining (ICS), a flow-cytometry-based assay that detects specific T-cell types (e.g. CD4+, CD8+) after cellular immunity is induced by external antigens. Cytokines included interferon (IFN)-γ, interleukin (IL)-2, tumour necrosis factor (TNF)-α, and IL-4.

### Outcomes

The co-primary endpoints were neutralising antibody titres (GMT and SCR) to SARS-CoV-2 parental strain (D614G) measured by live virus neutralisation assay (FRNT) at 2 weeks after the second vaccination. Comparison between groups was measured as GMT ratio and SCR difference, defined as the percentage of participants with ≥4-fold rise in live virus neutralising antibody titre from baseline to 2 weeks after the second vaccination.

Secondary immunogenicity endpoints included titres of SARS-CoV-2 RBD-binding IgG antibody measured by ELISA and neutralising antibody to SARS-CoV-2 measured by live virus neutralisation assay, at each timepoint post-vaccination, measured as GMT, GMT ratio, geometric mean fold rise (GMFR) from baseline, SCR and SCR difference. Additionally, cell-mediated responses for cytokines, measured by FluoroSpot and, for CD4+ and CD8+ T-cells, measured by ICS, after the second vaccination were considered. Safety endpoints (secondary endpoints) included: immediate unsolicited systemic reactions (within 30 minutes post-vaccination); solicited local and systemic AEs within 7 days post-vaccination; unsolicited AEs within 28 days post-vaccination; SAEs, MAAEs and AESIs during the whole study period.

The current interim analysis includes immunogenicity at 2 weeks and safety data (including immediate unsolicited systemic AEs, solicited local/systemic AEs, unsolicited AEs, SAEs, MAAEs, AEs leading to withdrawal, and AESIs) up to 4 weeks after the second vaccination (data cut-off 18 March 2022). Additionally, a median 2.5 months of safety follow-up after the last vaccination is included and contributed to the main safety data set (data cut-off 31 March 2022). Results for 6 and 12 months (durability of response and safety) are not yet available.

### Statistical analysis

The sample size for Cohort 1 was based on neutralisation assay results from the Phase 3 study of ChAdOx1-S^8^ and the Phase 1/2 study for GBP510/AS03^3^ to demonstrate superiority of GMT ratio and non-inferiority of SCR difference in terms of neutralising antibody titres. The planned sample size for Cohort 1 was 1950 (1300 test group, 650 control group) and that for Cohort 2 was 2040 (1700 test group, 340 control group); overall, this would ensure safety data were available for at least 3000 recipients of the two-dose GBP510/AS03 regimen. This sample size was based on FDA guidance for vaccine industry,^10^ and may allow the observance of at least one adverse event occurring at a frequency of 1 in 1,000. If the true AE rate was 0.1%, there would be a 95.03% probability of observing at least 1 AE in a test group comprising 3000 subjects.

All participants who received at least one dose of study vaccine were included in the safety set. The full analysis set comprised all participants who received at least 1 dose of the study vaccine and had valid pre- and at least one post-vaccination immunogenicity assessment results. The interim analysis of immunogenicity was primarily reported for the per-protocol population (baseline neutralising antibody titer was below LLOQ in wild-type virus neutralisation assay and completed vaccination schedule without SARS-CoV-2 infection with no major protocol deviations).

For the co-primary endpoints, adjusted post-vaccination GMT ratio estimate and 95% CIs were determined using analysis of covariance (ANCOVA) on log-transformed titres with treatment group and age (18–64 or ≥65 years) as factors, and baseline antibody level (titre) as covariate, while the 95% CIs for the difference in SCRs were calculated based on Chan and Zhang methodology. Superiority of post-vaccination GMT was demonstrated if the lower limit of the two-sided 95% CI for the ratio of post-vaccination GMTs at 2 weeks after the second study vaccination (GBP510/AS03 / ChAdOx1-S) was greater than 1. Non-inferiority of SCR was demonstrated if the lower limit of the two-sided 95% CI for the difference in the percentage of participants with a ≥4-fold rise from baseline in neutralisation antibody titre at 2 weeks after the second vaccination (GBP510/AS03 – ChAdOx1-S) exceeded the non-inferiority margin of –5%. For secondary immunogenicity endpoints, point estimates and 95% CI, or summary statistics (n, mean, SD, median, min and max), were presented for each treatment group. For safety endpoints, number and percentages of participants with at least one AE were presented by treatment group.

## RESULTS

### Characteristics of participants

Between 30 August 2021 and 10 February 2022, a total of 4036 participants (1956 in Cohort 1 and 2080 in Cohort 2) were randomised to receive GBP510/AS03 (n=3039) or ChAdOx1-S (n=997); Figure 1. After exclusion of 11 randomised individuals who did not receive vaccine, the safety dataset included 4025 subjects (3029 in the GBP510/AS03 group and 996 in the ChAdOx1-S group). Among Cohort 1 participants, 1887 (1259 in the GBP510/AS03 group and 628 in the ChAdOx1-S group) received at least 1 dose of the study vaccine and had valid pre- and at least one post-vaccination immunogenicity results (full analysis set). The per-protocol set for the interim analysis of immunogenicity included 1318 subjects (877 in the GBP510/AS03 group and 441 in the ChAdOx1-S group) (Figure S2).

**Figure 1.**
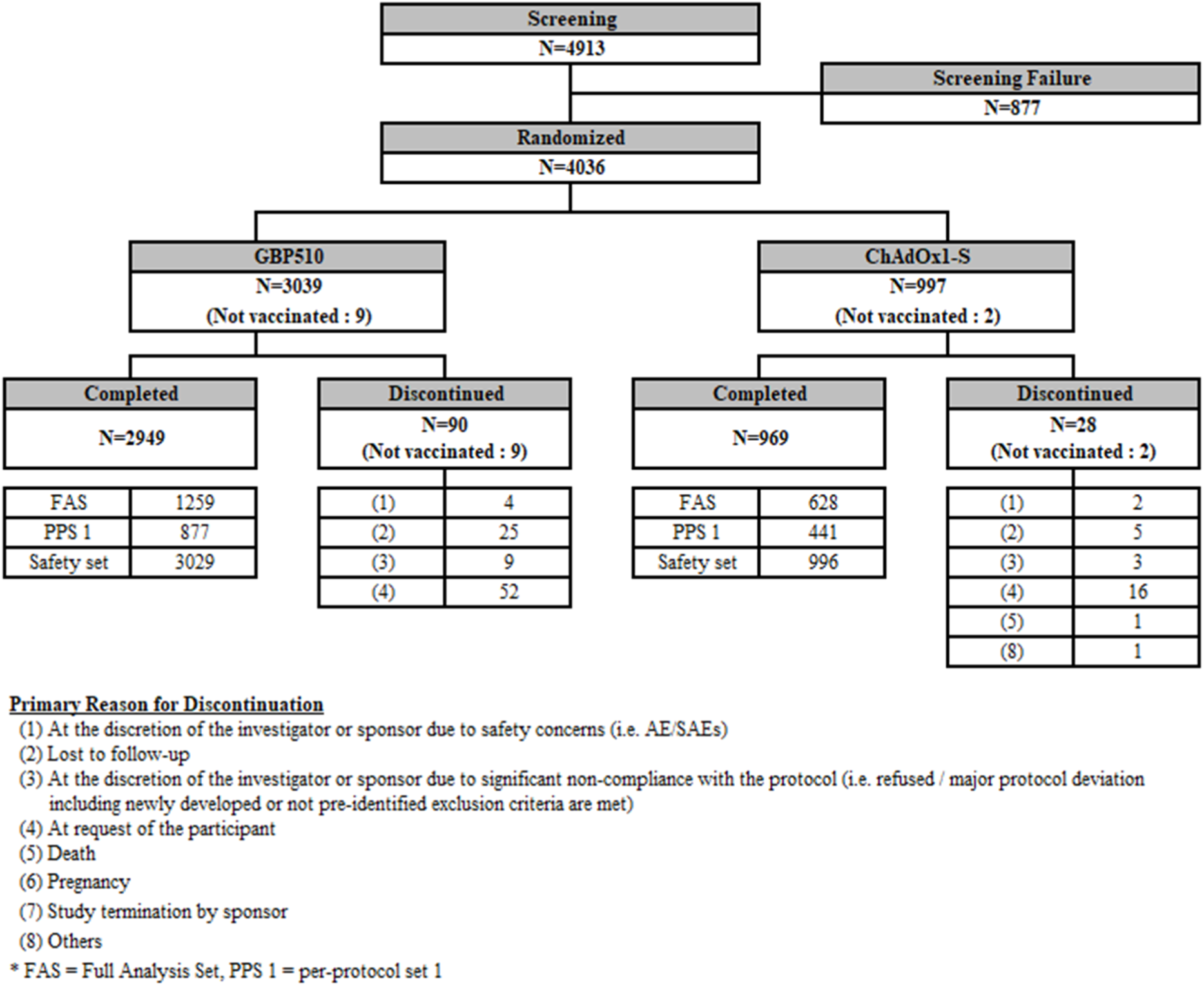
Study disposition.

The demographics and baseline characteristics of study participants are summarised in Table 1. Most subjects were Southeast Asian (81.47%). There were more males (59.12%) than females (40.88%). The mean age of subjects was 38.20±13.83 years; the mean age of the GBP510/AS03 group was slightly lower than that of the ChAdOx1-S group (37.83±13.84 versus 39.31±13.77, p=0.0034). However, the age strata distribution did not differ between the groups, with a total of 94.67% of subjects aged 18–64 years. The mean body mass index was 23.67±4.33 kg/m^2^.

**Table 1.** Demographics and baseline characteristics

|  | **GBP510/AS03**<br>**(N=3039)** | **ChAdOx1-S**<br>**(N=997)** | **Total**<br>**(N=4036)** |
| --- | --- | --- | --- |
| Age (years), mean ±SD | 37.83 ±13.84* | 39.31 ±13.77* | 38.20 ± 13.83 |
| 18–64 years, n (%) | 2878 (94.70) | 943 (94.58) | 3821 (94.67) |
| ≥65 years, n (%) | 161 (5.30) | 54 (5.42) | 215 (5.33) |
| Male/Female, n (%) |  |  |  |
| Male | 1814 (59.69) | 572 (57.37) | 2386 (59.12) |
| Female | 1225 (40.31) | 425 (42.63) | 1650 (40.88) |
| Race, n (%) |  |  |  |
| Asian | 2856 (93.98) | 931 (93.38) | 3787 (93.83) |
| Korean | 329 (10.83) | 168 (16.85) | 497 (12.31) |
| Southeast Asian | 2526 (83.12) | 762 (76.43) | 3288 (81.47) |
| Other Asian | 1 (0.03) | 1 (0.10) | 2 (0.05) |
| Caucasian | 177 (5.82) | 64 (6.42) | 241 (5.97) |
| Black | 0 | 0 | 0 |
| Hispanic | 0 | 0 | 0 |
| Other | 6 (0.20) | 2 (0.20) | 8 (0.20) |
| Body mass index (kg/m<sup>2</sup>), mean ±SD | 23.59 ±4.35 | 23.90 ±4.27 | 23.67 ±4.33 |
| Concomitant illness, n (%) | 486 (15.99) | 204 (20.46) | 690 (17.10) |
| Concomitant medication, n (%) | 1232 (40.54) | 411 (41.22) | 1643 (40.71) |

\* Between group p=0.0034.

### Immunogenicity outcomes

#### Live virus neutralisation assays (FRNT)

The co-primary endpoints were based on measurement of the neutralising antibody response (ND_50_ presented as IU/mL using the WHO standard) to SARS-CoV-2 using live virus neutralisation assays (FRNT). Post-vaccination GMTs were adjusted for age group (18–64 or ≥65 years) and baseline antibody level.

At baseline, there was no significant difference in GMT between the GBP510/AS03 group and the ChAdOx1-S group. At 4 weeks after the first vaccination, GMT (Figure 2), GMFR, and SCR were higher in the ChAdOx1-S group than in the GBP510/AS03 group, with between-group differences for the GMT ratio and SCR achieving statistical significance (Table 2).

**Figure 2.**
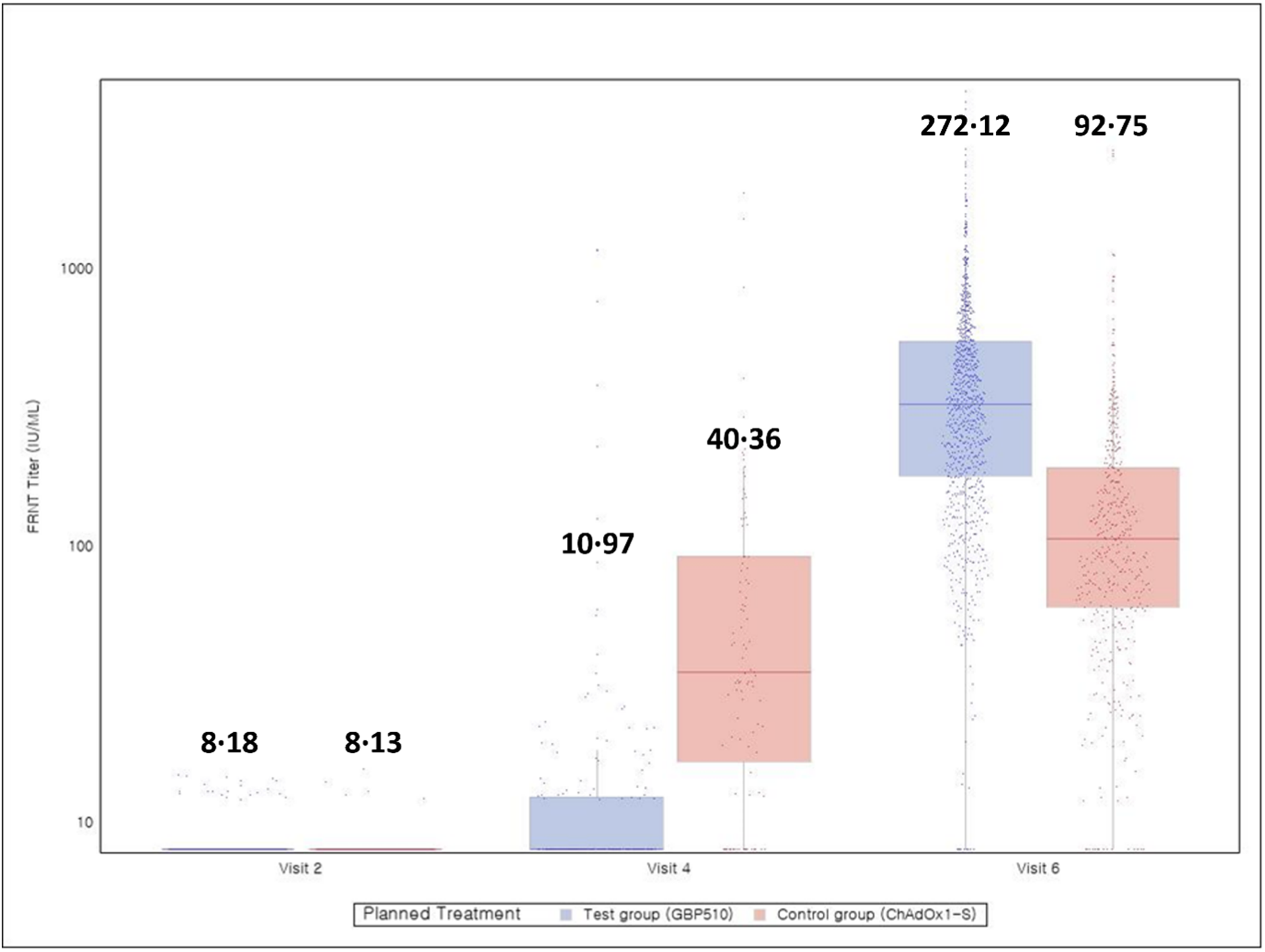
Boxplots and individual data for the natural logarithmic titre by live virus neutralisation assay at baseline (visit 2), 4 weeks after first vaccination (visit 4), and 2 weeks after second vaccination (visit 6) (per-protocol set)

**Table 2.**
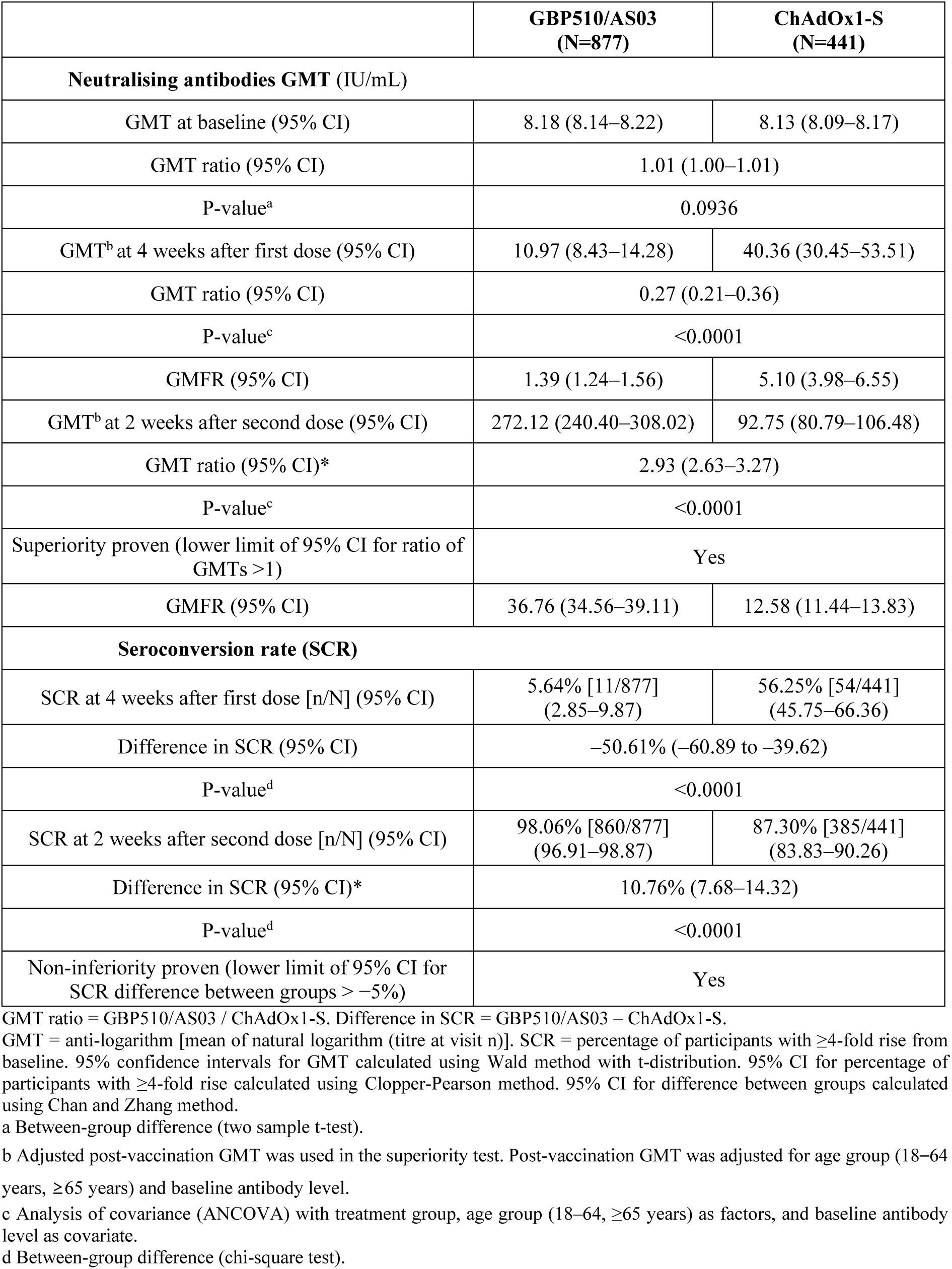
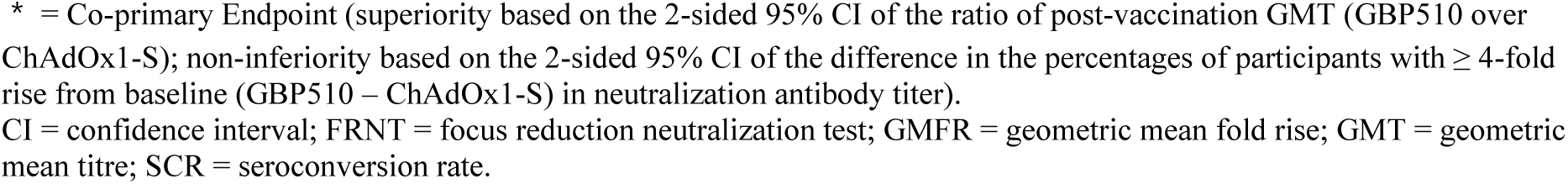
Immunogenicity assessment: neutralising antibody response to SARS-CoV-2 measured by live-virus neutralisation assays (FRNT). GMT, GMFR and SCR at 2 weeks after the second dose of vaccine were co-primary endpoints (per-protocol set)

|  | **GBP510/AS03**<br>**(N=877)** | **ChAdOx1-S**<br>**(N=441)** |
| --- | --- | --- |
| **Neutralising antibodies GMT (IU/mL)** |  |  |
| GMT at baseline (95% CI) | 8.18 (8.14–8.22) | 8.13 (8.09–8.17) |
| GMT ratio (95% CI) | 1.01 (1.00–1.01) |  |
| P-value<sup>a</sup> | 0.0936 |  |
| GMT<sup>b</sup> at 4 weeks after first dose (95% CI) | 10.97 (8.43–14.28) | 40.36 (30.45–53.51) |
| GMT ratio (95% CI) | 0.27 (0.21–0.36) |  |
| P-value<sup>c</sup> | <0.0001 |  |
| GMFR (95% CI) | 1.39 (1.24–1.56) | 5.10 (3.98–6.55) |
| GMT<sup>b</sup> at 2 weeks after second dose (95% CI) | 272.12 (240.40–308.02) | 92.75 (80.79–106.48) |
| GMT ratio (95% CI)* | 2.93 (2.63–3.27) |  |
| P-value<sup>c</sup> | <0.0001 |  |
| Superiority proven (lower limit of 95% CI for ratio of GMTs >1) | Yes |  |
| GMFR (95% CI) | 36.76 (34.56–39.11) | 12.58 (11.44–13.83) |
| **Seroconversion rate (SCR)** |  |  |
| SCR at 4 weeks after first dose [n/N] (95% CI) | 5.64% [11/877]<br>(2.85–9.87) | 56.25% [54/441]<br>(45.75–66.36) |
| Difference in SCR (95% CI) | –50.61% (–60.89 to –39.62) |  |
| P-value<sup>d</sup> | <0.0001 |  |
| SCR at 2 weeks after second dose [n/N] (95% CI) | 98.06% [860/877]<br>(96.91–98.87) | 87.30% [385/441]<br>(83.83–90.26) |
| Difference in SCR (95% CI)* | 10.76% (7.68–14.32) |  |
| P-value<sup>d</sup> | <0.0001 |  |
| Non-inferiority proven (lower limit of 95% CI for SCR difference between groups > –5%) | Yes |  |

GMT ratio = GBP510/AS03 / ChAdOx1-S. Difference in SCR = GBP510/AS03 – ChAdOx1-S.
GMT = anti-logarithm [mean of natural logarithm (titre at visit n)]. SCR = percentage of participants with ≥4-fold rise from baseline. 95% confidence intervals for GMT calculated using Wald method with t-distribution. 95% CI for percentage of participants with ≥4-fold rise calculated using Clopper-Pearson method. 95% CI for difference between groups calculated using Chan and Zhang method.
a Between-group difference (two sample t-test).
b Adjusted post-vaccination GMT was used in the superiority test. Post-vaccination GMT was adjusted for age group (18–64 years, ≥65 years) and baseline antibody level.
c Analysis of covariance (ANCOVA) with treatment group, age group (18–64, ≥65 years) as factors, and baseline antibody level as covariate.
d Between-group difference (chi-square test).
\* = Co-primary Endpoint (superiority based on the 2-sided 95% CI of the ratio of post-vaccination GMT (GBP510 over ChAdOx1-S); non-inferiority based on the 2-sided 95% CI of the difference in the percentages of participants with $\geq 4$ -fold rise from baseline (GBP510 – ChAdOx1-S) in neutralization antibody titer). CI = confidence interval; FRNT = focus reduction neutralization test; GMFR = geometric mean fold rise; GMT = geometric mean titre; SCR = seroconversion rate.

At 2 weeks after the second vaccination (when the co-primary endpoints were evaluated), GMT (Figure 2) and SCR were higher in the GBP510/AS03 group than in the ChAdOx1-S group, as follows. The adjusted GMT ratio (GBP510/AS03 / ChAdOx1-S) was 2.93 (95% CI 2.63–3.27), which was statistically significantly different from 1 (p<0.0001) and satisfying the hypothesis of superiority (lower limit of 95% CI greater than 1);. The between-group difference in the SCR was 10.76% (95% CI 7.68–14.32), satisfying the hypothesis of non-inferiority (lower limit of 95% CI greater than −5%);. The test for independence between SCR and group was statistically significant (p<0.0001). In addition, the GMFR (±SD) was also higher in the GBP510/AS03 group (36.76±2.54) than in the ChAdOx1 group (12.58±2.76) at 2 weeks after the second vaccination (Table 2).

In subgroup analyses, similar results were seen at 2 weeks after the second vaccination in younger (18–64 years) and older (≥65 years) age groups (Tables S3–S4), in male and female, and across ethnicities (Korean, Southeast Asian and Caucasian; Tables S5), and results in participants who were seropositive for neutralising antibody at baseline (≥LLOQ) were consistent with the seronegative population.

In addition, neutralising antibody GMTs at 2 weeks after the second vaccination (FRNT presented as ND_50_) were higher with GBP510/AS03 than with ChAdOx1-S against the Delta (B.1.617.2) variant (2644.2 vs 96.96; GMT ratio 27.27 [95% CI 18.88–39.39], p<0.0001), the Omicron BA.1 (B.1.1.529) variant (129.09 vs 12.27; GMT ratio 10.52 [95% CI 8.18–13.53], p<0.0001), and the Omicron BA.5 sub-lineage (61.61 vs 13.56; GMT ratio 4.54 [95% CI 3.51– 5.88], p<0.0001) in a subset of participants (Tables S6-S8).

#### Enzyme-linked immunosorbent assay (ELISA)

The GMTs of SARS-CoV-2 RBD-binding IgG antibody measured by ELISA at baseline were similar between the two groups with no significant difference. At both 4 weeks after the first vaccination, and at 2 weeks after the second vaccination, GMT, GMFR, and the SCR were higher in the GBP510 group than in the ChAdOx1-S group, with differences in the GMT ratio and SCR achieving statistical significance at both timepoints (both p<0.0001) (Table 3, Figure S3).

**Table 3.** Immunogenicity assessment: SARS-CoV-2 RBD-binding IgG antibody measured using ELISA (secondary endpoints; per-protocol set)

|  | **GBP510/AS03**<br>**(N=877)** | **ChAdOx1-S**<br>**(N=441)** |
| --- | --- | --- |
| **ELISA GMT (BAU/mL)** |  |  |
| GMT at baseline (95% CI) | 10.91 (10.51–11.32) | 10.79 (10.27–11.34) |
| GMT ratio (95% CI) | 1.01 (0.95–1.08) |  |
| P-value<sup>a</sup> | 0.7409 |  |
| GMT<sup>b</sup> at 4 weeks after first dose (95% CI) | 131.24 (117.90–146.09) | 91.91 (81.55–103.57) |
| GMT ratio (95% CI) | 1.43 (1.30–1.57) |  |
| P-value<sup>c</sup> | <0.0001 |  |
| GMFR (95% CI) | 15.74 (14.79–16.76) | 11.07 (10.08–12.15) |
| GMT<sup>b</sup> at 2 weeks after second dose (95% CI) | 2850.45<br>(2586.51–3141.32) | 215.67 (193.52–240.36) |
| GMT ratio (95% CI) | 13.22 (12.13–14.40) |  |
| P-value<sup>c</sup> | <0.0001 |  |
| GMFR (95% CI) | 296.10 (278.11–315.24) | 23.02 (21.14–25.06) |
| **Seroconversion rate (SCR)** |  |  |
| SCR at 4 weeks after first dose [n/N] (95% CI) | 92.47% [811/877]<br>(90.52–94.13) | 84.81% [374/441]<br>(81.11–88.03) |
| Difference in SCR (95% CI) | 7.67 (3.89–11.44) |  |
| P-value<sup>d</sup> | <0.0001 |  |
| SCR at 2 weeks after second dose [n/N] (95% CI) | 99.54% [873/877]<br>(98.84–99.88) | 96.83% [427/441]<br>(94.73–98.25) |
| Difference in SCR (95% CI) | 2.72 (1.02–4.41) |  |
| P-value<sup>d</sup> | <0.0001 |  |

GMT ratio = GBP510/AS03 / ChAdOx1-S. Difference in SCR = GBP510/AS03 – ChAdOx1-S.
GMT = anti-logarithm [mean of natural logarithm (titre at visit n)]. GMFR = anti-logarithm [mean of natural logarithm (titre at visit n/titre at baseline)]. SCR = percentage of participants with ≥4-fold rise from baseline. 95% confidence intervals for GMT and GMFR calculated using Wald method with t-distribution. 95% CI for percentage of participants with ≥4-fold rise calculated using Clopper-Pearson method. 95% CI for difference between groups for participants with ≥4-fold rise using Wald method.
a Between-group difference (two sample t-test).
b Post-vaccination GMT was adjusted for age group (18–64 years, ≥65 years) and baseline antibody level.
c ANCOVA with treatment group, age group (18–64, ≥65 years) as factors, and baseline antibody level as covariate.
d Between-group difference (chi-square test).
ANCOVA = analysis of covariance; CI = confidence interval; ELISA = enzyme-linked immunosorbent assay; GMFR = geometric mean fold rise; GMT = geometric mean titre; RBD = receptor-binding domain.

#### Cell-mediated immune response

The cell-mediated immune response was evaluated in a subset of Cohort 1 participants. At 2 weeks after the second vaccination, the median cell count for IFNγ, TNF-α, and IL-2 increased from baseline in both groups based on FluoroSpot assays (Table 4, Figures S4–S6). In particular, noticeable increases in TNF-α response (from median 23.00 at baseline to 52.00 spot forming cell (SFC) /2.5×10^5^ peripheral blood mononuclear cells (PBMCs)) and IL-2 response (from 24.00 at baseline to 57.00 SFC/2.5×10^5^ PBMCs) were seen in the GBP510/AS03 group. No change in IL-4 was seen in either group (Table 4, Figure S7).

**Table 4.**
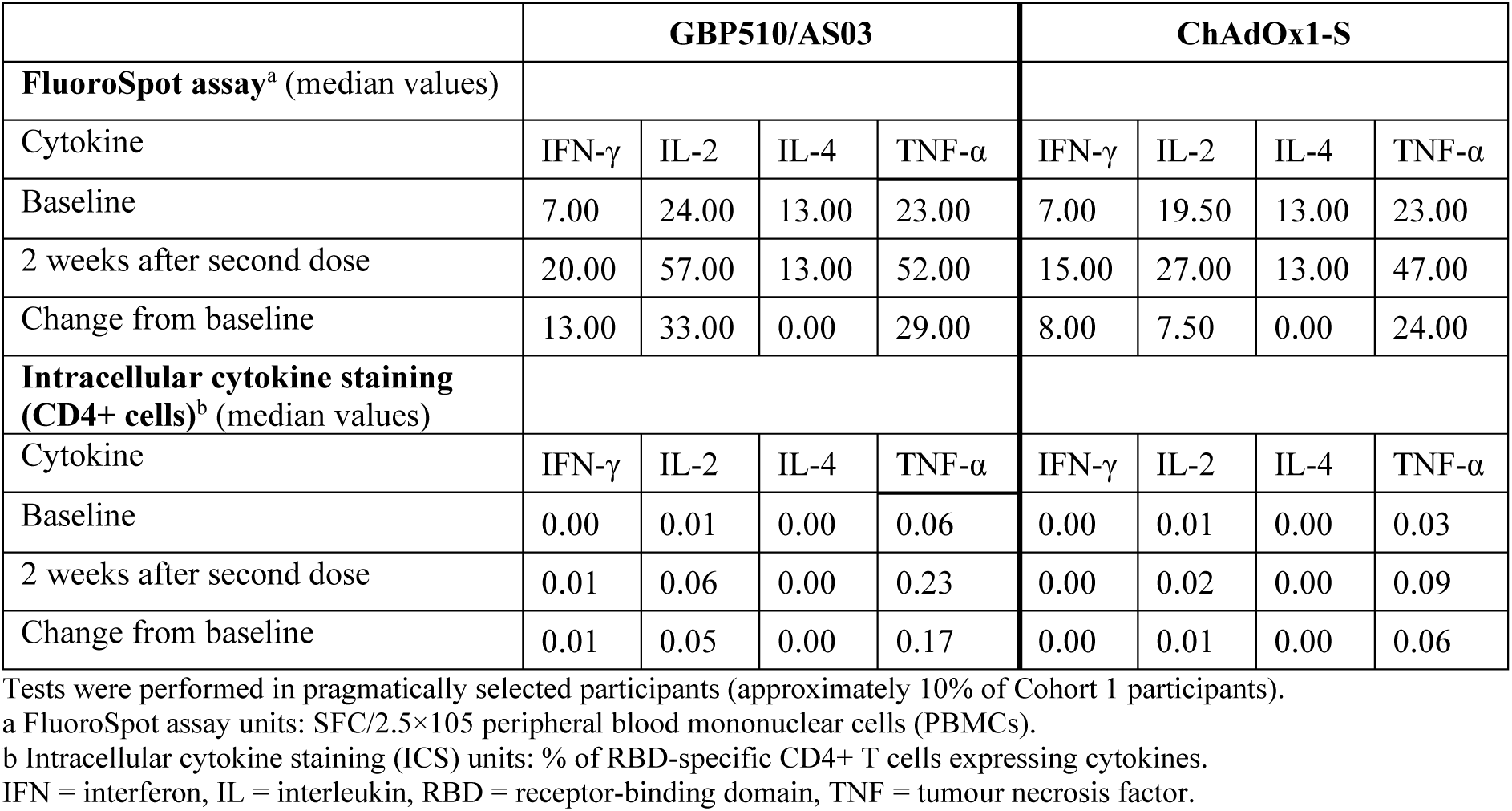
Cell-mediated immune response based on FluoroSpot assay (quantification of cytokine-secreting cells) and intracellular cytokine staining (for CD4+ T cells expressing cytokines)

In ICS assessments of CD4+ T cells, some response was observed for TNFα and IL-2 in the GBP510/AS03 group (Table 4, Figures S8–S11). No response was observed in CD8+ T cells in either group (Table S9).

### Safety outcomes

Of 3029 subjects in the GBP510/AS03 group and 996 subjects in the ChAdOx1-S group analysed for the Safety Set, immediate systemic AEs (occurring within 30 minutes) after any vaccination were reported by six subjects (0.20%) in the GBP510/AS03 group and in no subjects in the ChAdOx1-S group.

Solicited local AEs after any vaccination had a higher incidence in the GBP510/AS03 group (56.69% of subjects) than in the ChAdOx1-S group (49.20% of subjects; p<0.0001). Injection site pain was the most common solicited local AE in both groups, reported by 55.69% of subjects in the GBP510/AS03 group and 48.80% of subjects in the ChAdOx1-S group; injection site redness was reported by 5.25% and 2.01% of subjects, and injection site swelling by 5.55% and 1.20% of subjects (Figure 3).

**Figure 3.**
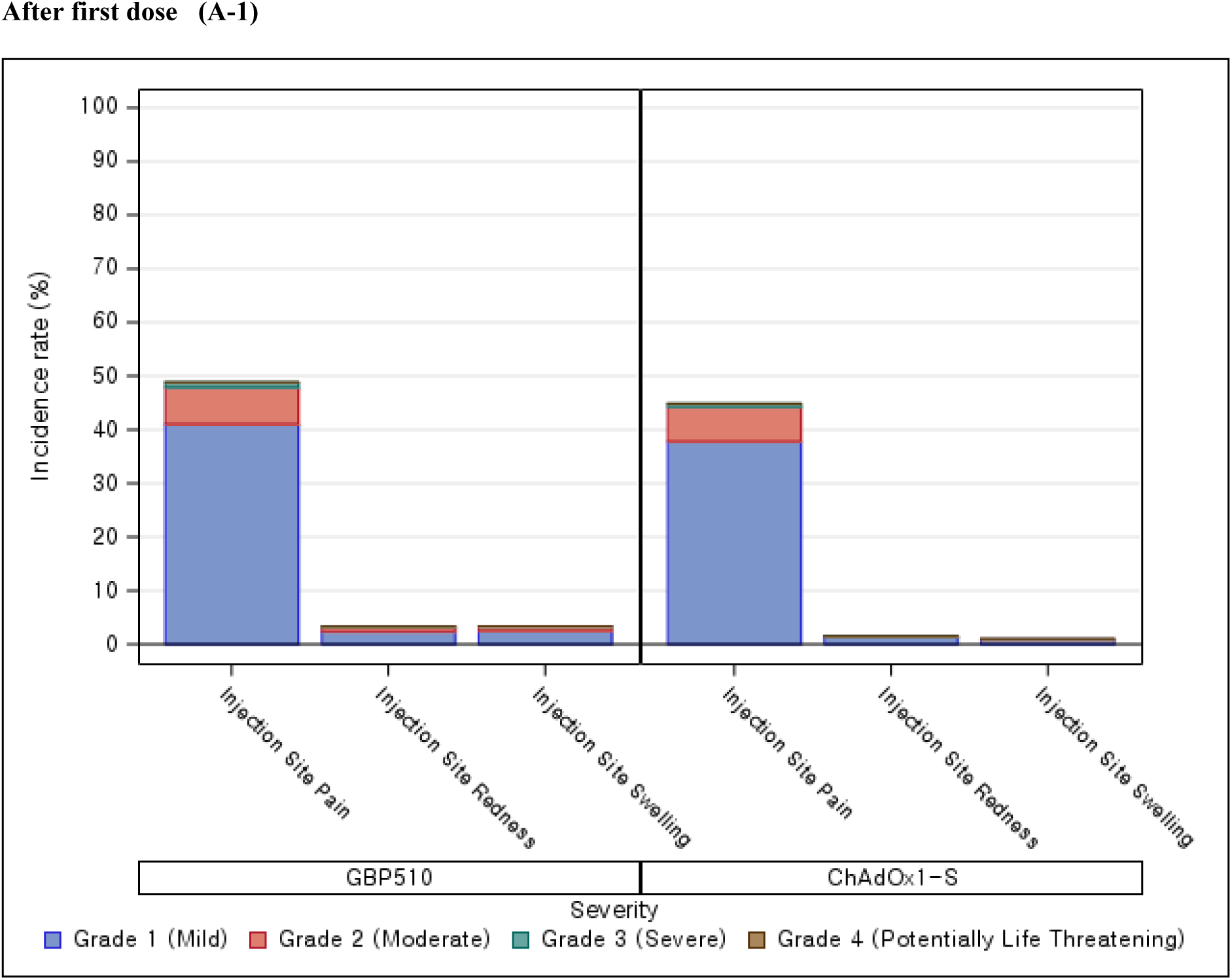
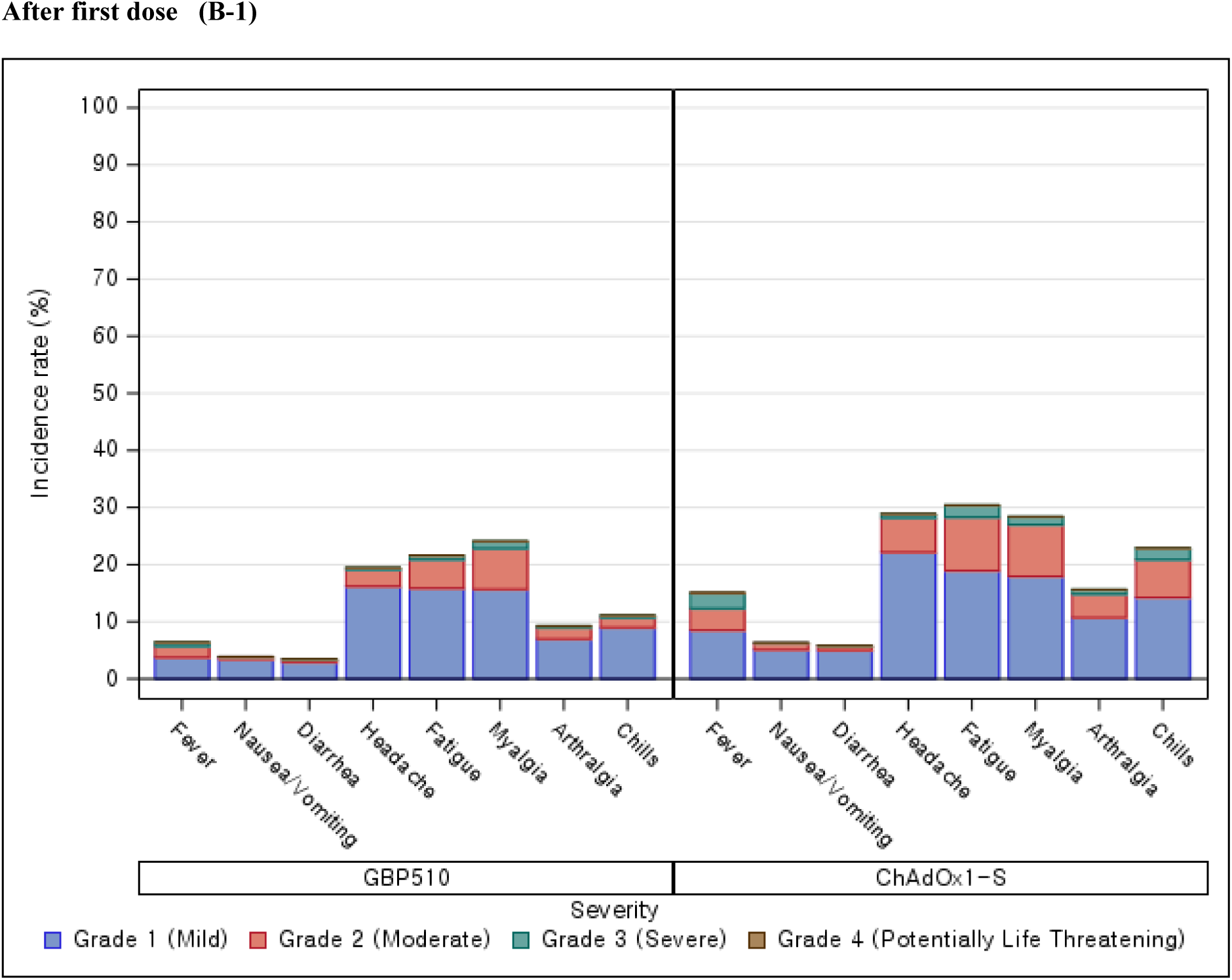
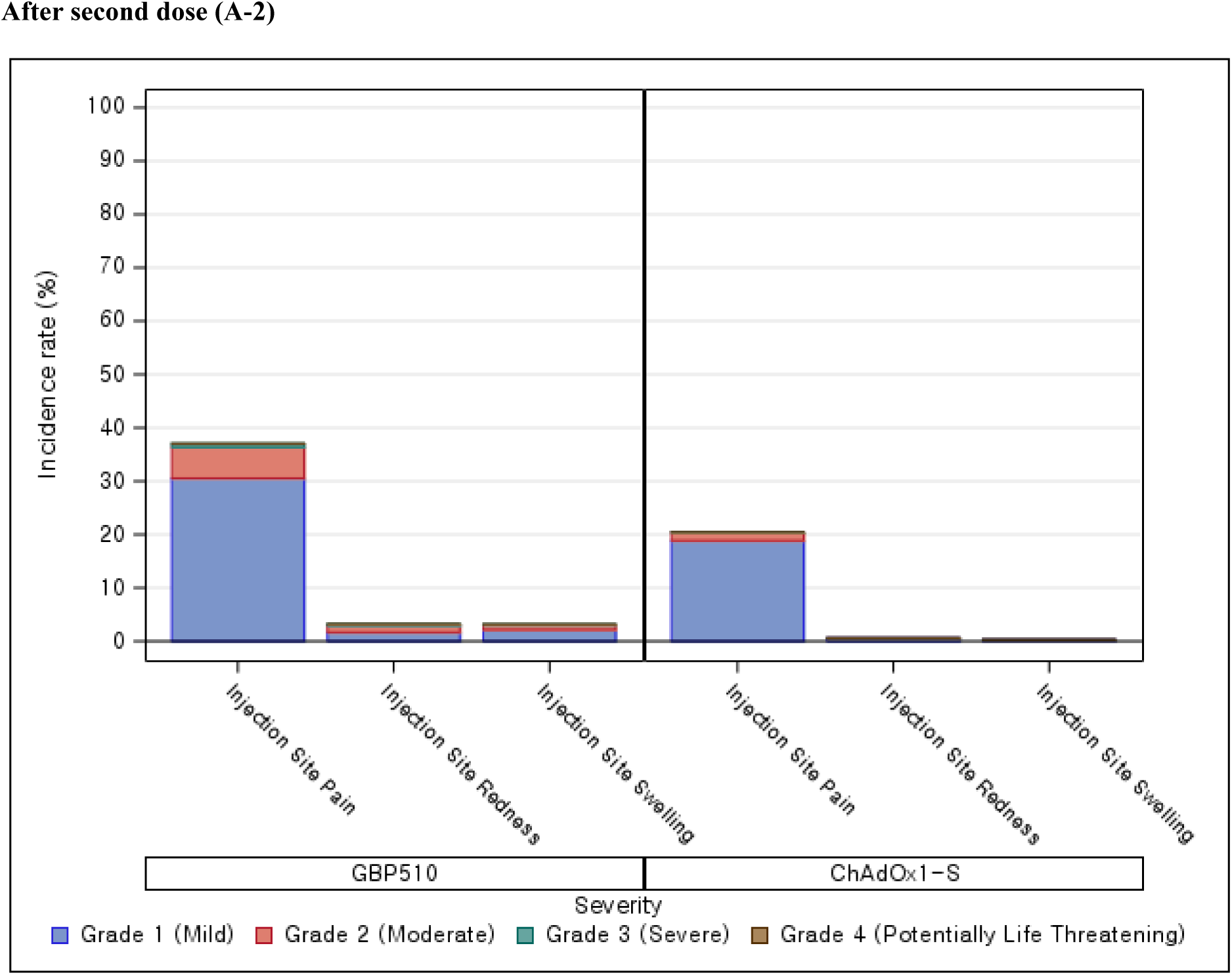
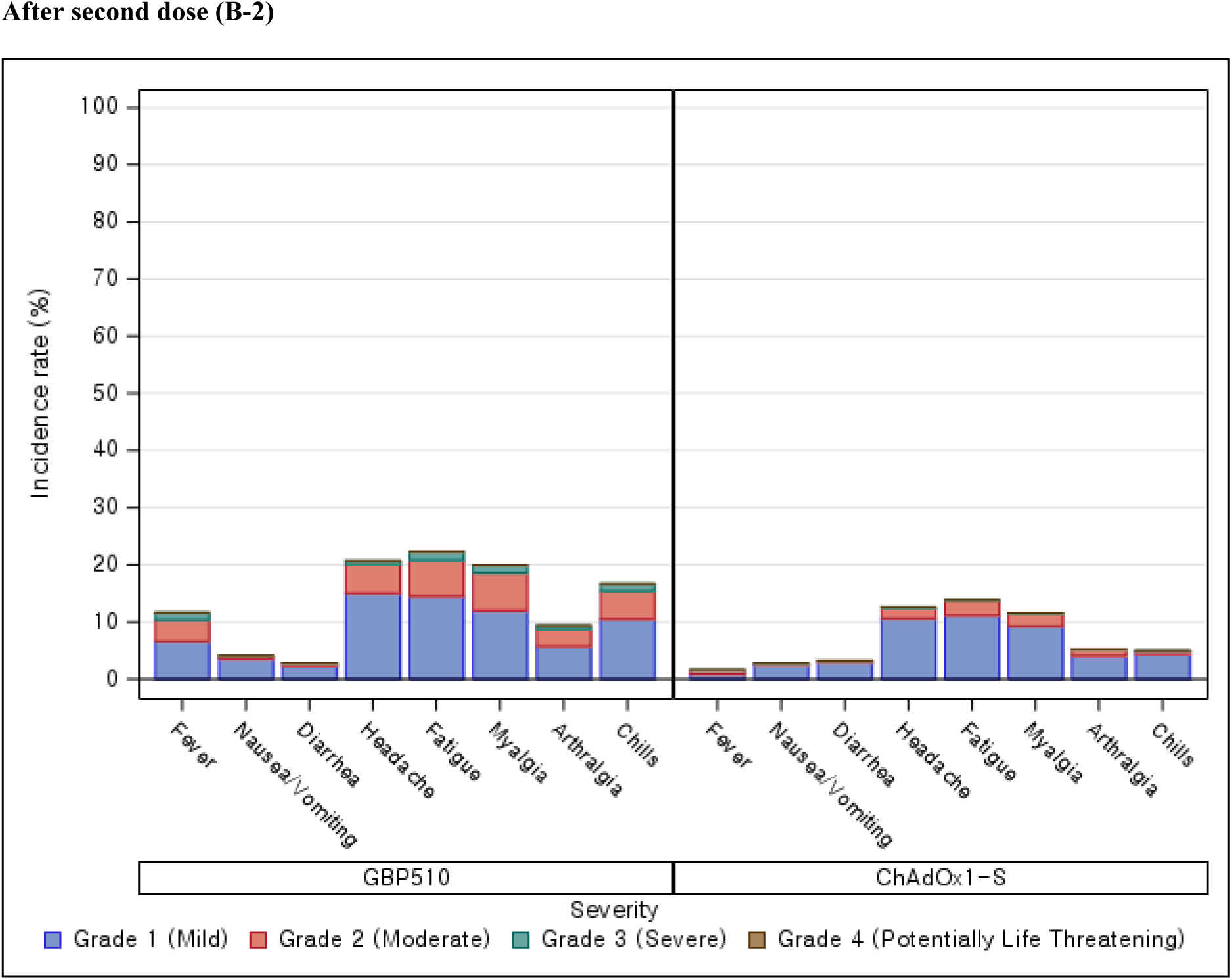
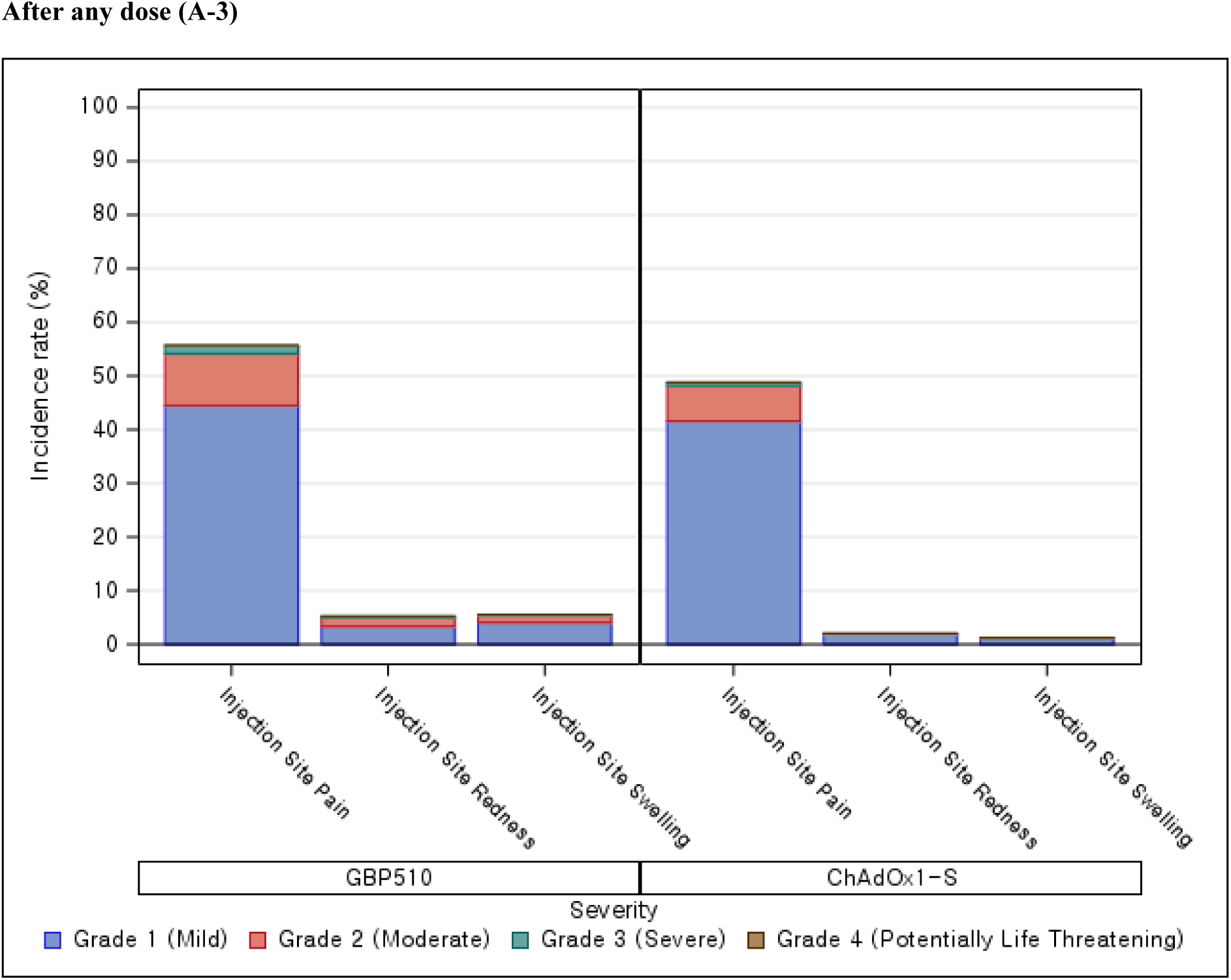
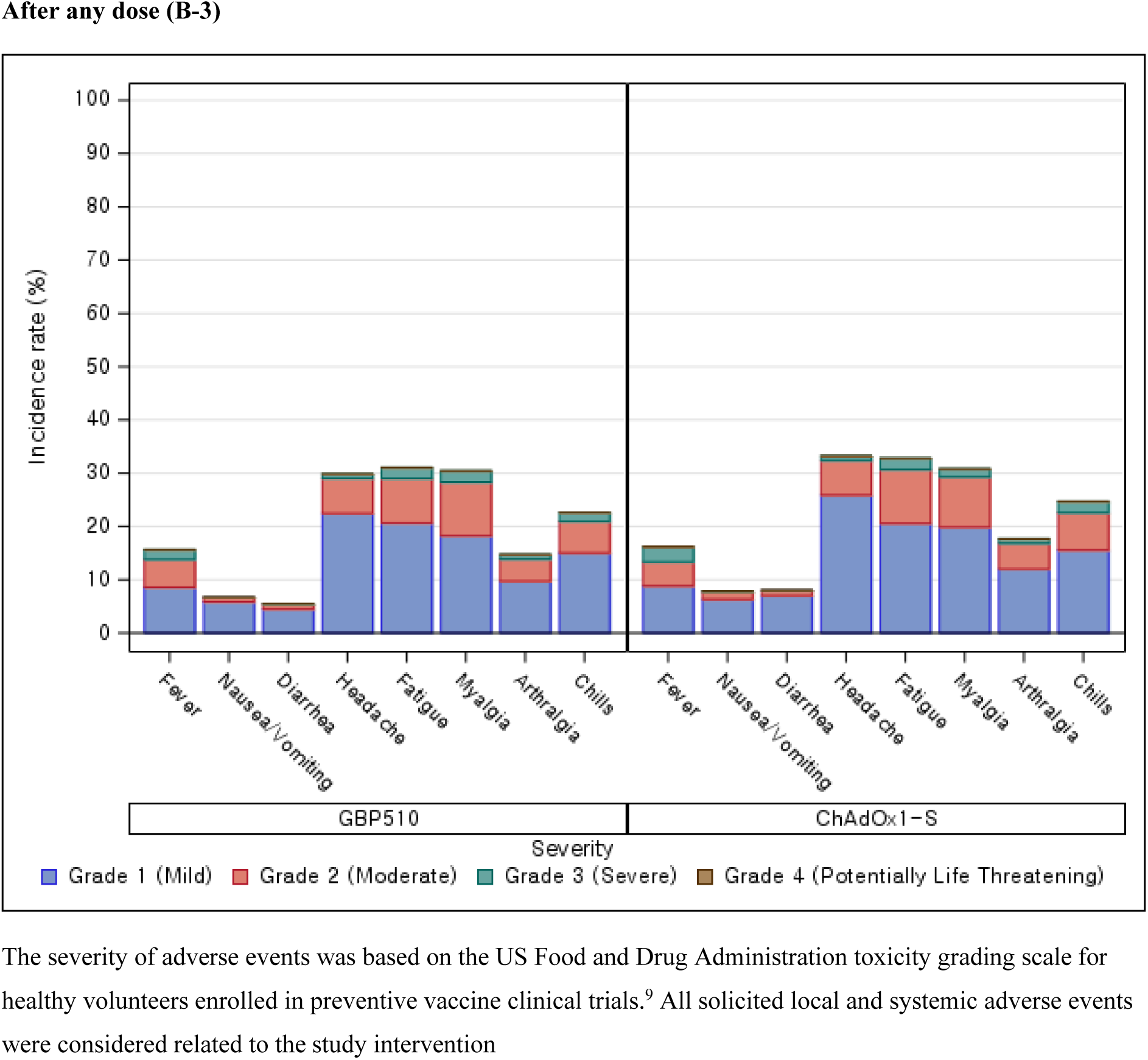
Incidence rates for solicited local (A) and systemic (B) adverse events within 7 days after each-dose vaccination.

Solicited systemic AEs after any vaccination were reported by 51.21% of subjects in the GBP510/AS03 group and 53.51% of subjects in the ChAdOx1-S group (p=0.2058). Headache (29.88% vs 33.23%), fatigue (31.07% vs 32.83%), and myalgia (30.47% vs 30.82%) were the most commonly reported solicited systemic AEs in both groups after any vaccination . In the GBP510/AS03 group, the frequency of solicited systemic AEs was similar after the first and second doses, while in the ChAdOx1-S group the frequency was higher after the first dose than after the second dose (Figure 3).

Most solicited local and systemic AEs were grade 1 (mild) or 2 (moderate) in severity and lasted a mean of 1.3 to 3.1 days for both groups (Figure 3, Tables S10 and S11). The proportion of participants who reported grade 3 (severe) solicited local AE after any vaccination was 1.85% in the GBP510/AS03 group and 0.60% in the ChAdOx1-S group. Grade 3 solicited systemic AEs were reported by 5.28% of participants in GBP510/AS03 group and 6.12% in ChAdOx1-S group. No grade 4 (potentially life threatening) solicited AEs were reported in either group.

The proportions of subjects aged 18–64 years with solicited local/systemic AEs were 56.70%/51.41% in the GBP510/AS03 group and 49.42%/53.67% in the ChAdOx1-S group, and in subjects aged ≥65 years were 56.33%/47.47% and 45.45%/50.91%, respectively. The proportion of Korean subjects reporting solicited local/systemic AEs (96.94%/93.58% in the GBP510/AS03 group and 85.12%/92.86% in the ChAdOx1-S group) was higher than seen among Southeast Asian subjects (49.90%/43.95% and 39.68%/43.76%, respectively).

Unsolicited AEs occurring within 28 days after any vaccination were reported by 13.27% of subjects in the GBP510/AS03 group and 14.56% of subjects in the ChAdOx1-S group (p=0.3040), including 3.50% and 4.12%, respectively, of unsolicited adverse drug reactions (ADRs) (p=0.3679) (Table S12). The most commonly reported unsolicited AEs by System Organ Class (SOC) were ‘Infections and infestations’ in both the GBP510/AS03 (5.91% of subjects) and the ChAdOx1-S (5.92% of subjects) groups, which were predominantly due to COVID-19 infections. Most unsolicited AEs after any vaccination were grade 1 (mild) or 2 (moderate) in severity. Unsolicited AEs of grade ≥3 severity occurred in 0.66% of subjects in the GBP510/AS03 group and 1.20% in the ChAdOx1-S group (p=0.0932).

MAAEs were reported for 7.49% of subjects in the GBP510/AS03 group and 8.43% of subjects in the ChAdOx1-S group, including 0.63% and 1.00% of subjects, respectively, with MAADRs (Table 5).

**Table 5.**
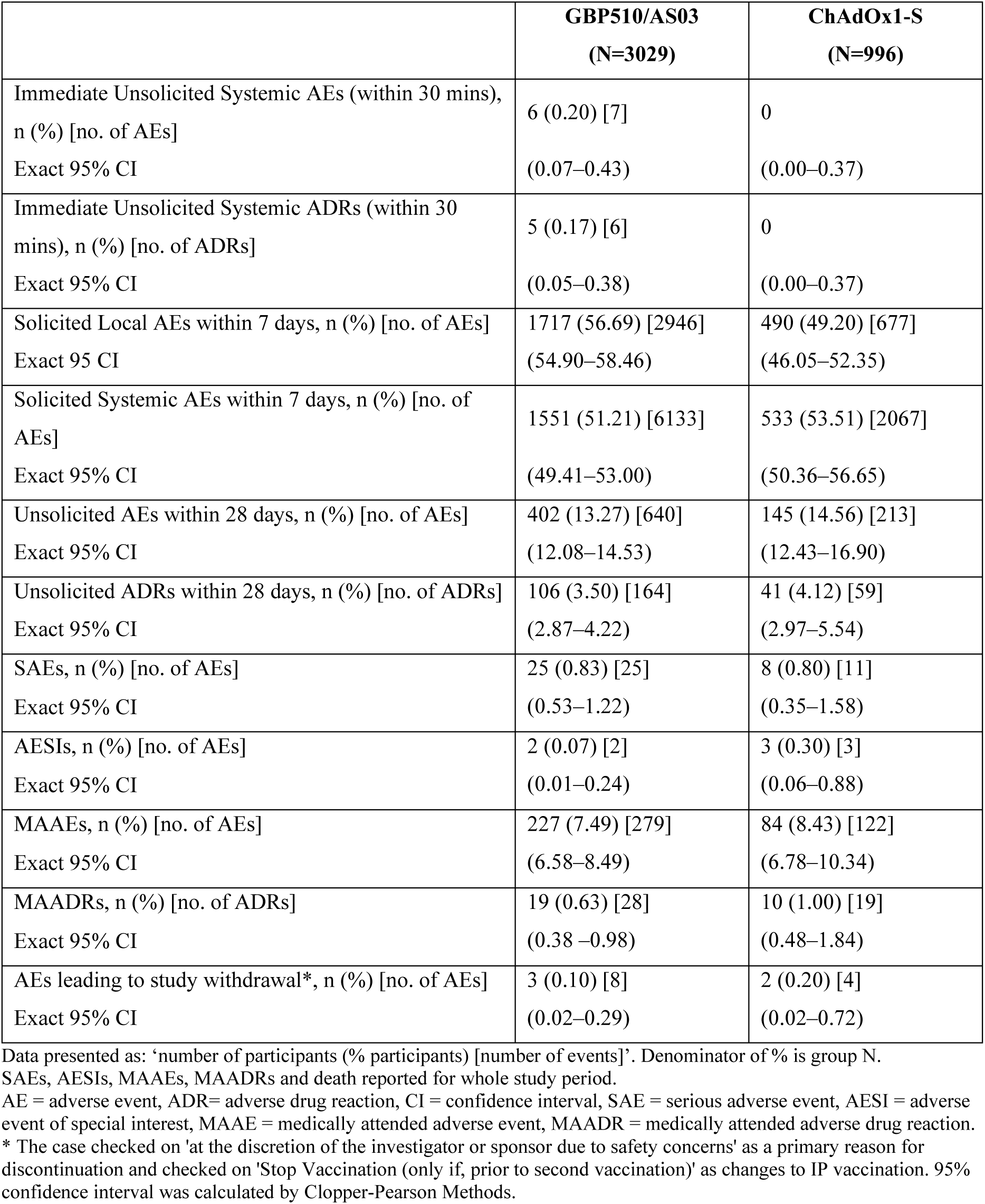
Overall adverse events (after any study vaccination) up to median 2.5 months after second vaccination (safety dataset)

|  | **GBP510/AS03**<br>**(N=3029)** | **ChAdOx1-S**<br>**(N=996)** |
| --- | --- | --- |
| Immediate Unsolicited Systemic AEs (within 30 mins),<br>n (%) [no. of AEs]<br>Exact 95% CI | 6 (0.20) [7]<br>(0.07–0.43) | 0<br>(0.00–0.37) |
| Immediate Unsolicited Systemic ADRs (within 30<br>mins), n (%) [no. of ADRs]<br>Exact 95% CI | 5 (0.17) [6]<br>(0.05–0.38) | 0<br>(0.00–0.37) |
| Solicited Local AEs within 7 days, n (%) [no. of AEs]<br>Exact 95% CI | 1717 (56.69) [2946]<br>(54.90–58.46) | 490 (49.20) [677]<br>(46.05–52.35) |
| Solicited Systemic AEs within 7 days, n (%) [no. of<br>AEs]<br>Exact 95% CI | 1551 (51.21) [6133]<br>(49.41–53.00) | 533 (53.51) [2067]<br>(50.36–56.65) |
| Unsolicited AEs within 28 days, n (%) [no. of AEs]<br>Exact 95% CI | 402 (13.27) [640]<br>(12.08–14.53) | 145 (14.56) [213]<br>(12.43–16.90) |
| Unsolicited ADRs within 28 days, n (%) [no. of ADRs]<br>Exact 95% CI | 106 (3.50) [164]<br>(2.87–4.22) | 41 (4.12) [59]<br>(2.97–5.54) |
| SAEs, n (%) [no. of AEs]<br>Exact 95% CI | 25 (0.83) [25]<br>(0.53–1.22) | 8 (0.80) [11]<br>(0.35–1.58) |
| AESIs, n (%) [no. of AEs]<br>Exact 95% CI | 2 (0.07) [2]<br>(0.01–0.24) | 3 (0.30) [3]<br>(0.06–0.88) |
| MAAEs, n (%) [no. of AEs]<br>Exact 95% CI | 227 (7.49) [279]<br>(6.58–8.49) | 84 (8.43) [122]<br>(6.78–10.34) |
| MAADRs, n (%) [no. of ADRs]<br>Exact 95% CI | 19 (0.63) [28]<br>(0.38–0.98) | 10 (1.00) [19]<br>(0.48–1.84) |
| AEs leading to study withdrawal*, n (%) [no. of AEs]<br>Exact 95% CI | 3 (0.10) [8]<br>(0.02–0.29) | 2 (0.20) [4]<br>(0.02–0.72) |

Data presented as: 'number of participants (% participants) [number of events]'. Denominator of % is group N.
SAEs, AESIs, MAAEs, MAADRs and death reported for whole study period.
AE = adverse event, ADR= adverse drug reaction, CI = confidence interval, SAE = serious adverse event, AESI = adverse event of special interest, MAAE = medically attended adverse event, MAADR = medically attended adverse drug reaction.
\* The case checked on 'at the discretion of the investigator or sponsor due to safety concerns' as a primary reason for discontinuation and checked on 'Stop Vaccination (only if, prior to second vaccination)' as changes to IP vaccination. 95% confidence interval was calculated by Clopper-Pearson Methods.

SAEs were reported for 0.83% of subjects in the GBP510/AS03 group and 0.80% in the ChAdOx1-S group. One event of rapidly progressive glomerulonephritis in the GBP510/AS03 group was assessed as a suspected unexpected serious adverse reaction (SUSAR) and AESI (Table 5). As a conservative judgement, the investigator reported it as a vaccine-related event due to the temporal relationship with GBP510/AS03 administration, and because a causal relationship with GBP510/AS03 could not be fully excluded. However, the clinical presentation and available laboratory findings do not provide evidence to establish a possible autoimmune aetiology for this event.

A total of 5 AESIs were reported; 2 in the GBP510/AS03 group (acute kidney injury and rapidly progressive glomerulonephritis) and 3 in the ChAdOx1-S group (acute pancreatitis, anaphylactic reaction, and psoriasis), of which only rapidly progressive glomerulonephritis was reported as a vaccine-related event.

One death occurred in the GBP510/AS03 group (brain neoplasm) and one death in ChAdOx1-S group (cardiorespiratory failure), with neither considered to be related to the study vaccines.

Five pregnancies were reported in the GBP510/AS03 group and two in the ChAdOx1-S group; the pregnancies were ongoing at the data lock point of the analysis, without any abnormal outcomes or SAEs reported during the reporting period. Three subjects in the GBP510/AS03 group and two in the ChAdOx1-S withdrew due to AEs, with one subject in the GBP510/AS03 group reported to have vaccine-related AEs (urticaria rash, injection site pain, headache, fatigue, myalgia, and arthralgia), all of which recovered/were resolved.

There were 336 virologically-confirmed (by PCR) or suspected (diagnosed with rapid antigen test) COVID-19 cases, 238 cases (7.86%) in the GBP510/AS03 group and 98 (9.84%) in the ChAdOx1-S group (Tables S13 and S14). All COVID-19 cases were either non-severe or asymptomatic, as per WHO criteria, thus, they were not suspected as vaccine-associated enhanced disease/vaccine-associated enhanced respiratory disease. However, COVID-19 was classified as an SAE in 0.33% of subjects in the GBP510/AS03 group and 0.40% in the ChAdOx1-S group, with the seriousness criterion being hospitalization. COVID-19 infected patients in some countries (e.g., Thailand) were required, under local regulations, to be monitored or treated in hospital regardless of their severity of symptoms or medical history; these were subsequently considered as serious COVID-19 cases.

## DISCUSSION

This interim analysis found that the immune response induced by two doses of GBP510/AS03 in seronegative adults was superior compared to ChAdOx1-S with respect to GMTs, and non-inferior with respect to SCRs, in terms of neutralising antibody response against the parental D614G strain of SARS-CoV-2 at 2 weeks after the second vaccination (primary endpoints). ELISA assessments of SARS-CoV-2 RBD-binding IgG antibody also indicated a higher immune response with GBP510/AS03 than with ChAdOx1-S at 2 weeks after the second vaccination.

Higher neutralising antibody responses were seen with GBP510/AS03 against the parental strain regardless of age or gender. In addition, a trend towards higher neutralising antibody responses against the Delta and Omicron variants was observed with GBP510/AS03 in comparison with ChAdOx-1S in a subset of participants.

Of note, when an additional analysis was performed in the subset to observe the trend in neutralising antibody response after interim analysis by FRNT, the GMT ratio of the two groups at 4 weeks after the second vaccination showed a similar trend to that seen at 2 weeks after the second vaccination (GMT ratio: 3.01 [95% CI 2.30–3.94]), whereas SCR in the ChAdOx1-S group showed a noticeable decrease compared to the GBP510/AS03 group (SCR: 94.74% [180/190 subjects; GBP510/AS03] and 68.18% [60/88 subjects; ChAdOx1-S]).

GBP510/AS03 had previously been found to be highly immunogenic in a phase 1/2 study.^3^ In that study, two-dose vaccination with GBP510 (25 μg RBD per dose)/AS03 induced high neutralising antibody titres measured by wild-type virus assay (plaque-reduction neutralisation test), with an increase in GMT from 4.33 at baseline to 861 IU/mL at 2 weeks after the second dose. Direct comparison with the current study is not possible because a different assay method (FRNT) was used; however, a high response was also seen in our study (GMT increased from 8.18 to 272.12 IU/mL). In the phase 1/2 study, a high geometric mean concentration of IgG antibody measured by ELISA was seen at 2 weeks after the second dose (2599 BAU/mL),^3^ which is consistent with the level observed at that timepoint in our study (2850.45 BAU/mL).

A noticeable increase in antibody responses after the second dose was observed compared to the first dose in both the phase 1/2^3^ and this phase 3 study, which may be due to some CD4+ T-cell response that was observed by ICS assessments after GBP510/AS03 vaccination. With CD4+ T-cell help, activated B cells initiate a germinal centre reaction, resulting in the generation of high-affinity memory B-cells and plasma cells, which are differentiated to antibody-secreting cells and provide overlapping layers of protection.^11, 12^ A superior antigen-specific CD4+ T-cell response was also reported with AS03-adjuvantation in a randomised influenza vaccine trial.^13^

GBP510/AS03 showed a clinically acceptable safety profile, and no safety concerns were noted during the study up to a median 2.5 months after the second vaccination. The incidence rates of unsolicited AEs and SAEs were similar between the study groups. Reactogenicity after any vaccination was higher with GBP510/AS03 than with ChAdOx1-S in terms of local solicited AEs, especially after the second vaccination, but was similar to ChAdOx1-S in terms of systemic solicited AEs (being lower after the first vaccination with GBP510/AS03, but higher after the second one, compared with ChAdOx1-S). Both local and systemic solicited AEs were mostly mild or moderate in intensity in both groups.

In the current study, GBP510/AS03 showed lower reactogenicity than that observed in the phase 1/2 study,^3^ which could be explained by a difference in study populations. Higher rates of solicited AEs were observed in Korean participants compared with Southeast Asians (who constituted the majority of study participants). The rates seen in Korean participants were consistent with those seen in the phase 1/2 study, which included only Korean subjects.^3^

One of the main limitations of the data generated in this study is that the absence of vaccine efficacy data, and the lack of established correlates of protection for COVID-19 vaccines, make it difficult to extrapolate the results of the study. Although thresholds for immune correlates of protection based on antibody levels or functional activity have not yet been established, COVID-19 vaccine studies that have reported vaccine efficacy have shown robust correlations between antibody responses (neutralising antibody or binding antibody titres) and vaccine efficacy, despite the use of different assays, endpoints, and study populations.^4, 14–16^ Another limitation is the small number of elderly participants due to high vaccination coverage in the elderly at the time of enrolment; however, the available results suggest that the immunogenicity of GBP510/AS03 against the parental strain was consistent across all age strata. The study included mostly Asian subjects, and did not include individuals with severe or unstable comorbid conditions, those who were immunocompromised, or pregnant or lactating women, and post-authorisation studies are needed to collect safety data in these populations. The main immunogenicity results from the study are for the D614 strain, whereas the current epidemiological context is dominated by Omicron subvariants; however, analysis in a subset of participants provided some evidence of neutralising antibody responses against Omicron BA.1 and BA.5 after vaccination with GBP510/AS03. Furthermore, additional studies are ongoing to evaluate the immunogenicity and safety of homologous and heterologous booster vaccination with GBP510/AS03. Finally, safety data for this interim analysis were only available up to a median 2.5 months after the second vaccination; however, the study is ongoing and additional data up to 12 months of follow-up will become available in due course.

In conclusion, this interim analysis found that two-dose vaccination with GBP510/AS03 induced stronger neutralising antibody immune responses compared with ChAdOx1-S against the ancestral D614G strain at 2 weeks after the second vaccination, and had a clinically acceptable safety profile.

## Contributors

HJC, HK, JHR, SJL, HKP, YWP, YYL, LC, FR, MC,TB, FS, PW, AP, and SGK contributed to the conception and design of this study. SS and PT contributed to the project administration. WSC, JYH, JSL, DSJ, SWK, KHP, JSE, SJJ, JL, KTK, HJC, JWS, YKK, HJC, BWY, IJJ, EJK, JC, EA, SN,WR, PM, RC, LQ, OK, TY, VK, AB, OG, OB, MP, CT, SC, SS, MW, THS, TH, DS, and MRC were the principal investigators of the study site. WSC, JYH, JSL, DSJ, SWK, KHP, JSE, SJJ, JL, KTK, HJC, JWS, YKK, BWY, IJJ, EJK, JYS, HJC, JC, EA, SN,WR, PM, RC, LQ, OK, TY, VK, AB, OG, OB, MP, CT, SC, SS, MW, THS, TH, DS, and MRC contributed to the acquisition of the clinical and laboratory data. BS and SS contributed to the resources of the study. SD contributed to the clinical operation. SS and MS contributed to the supervision of the study. JYS, SJJ, YKK, BWY, IJJ, PW, FR, MC,TB, and HJC contributed to the interpretation of the data. JYS and HJC had full access to and verify all of the data in the study and take responsibility for the integrity of the data. SS, ND, and SD contributed to the review and edit of the manuscript. JYS and HJC prepared the manuscript and had responsibility for the decision to submit the manuscript. All authors reviewed the manuscript for intellectual content and approved the final draft for submission. All authors have full access to all the data in the study and accept responsibility to submit for publication. All authors critically reviewed and approved the final version.

## Data sharing

Individual participant data will be made available when the trial is complete upon requests directed to the corresponding author. Proposals will be reviewed and approved by the sponsor, investigators, and collaborators on the basis of scientific merit. After approval of a proposal, data can be shared through a secure online platform.

## Data Availability

All authors reviewed the manuscript for intellectual content and approved the final draft for submission. All authors have full access to all the data in the study and accept responsibility to submit for publication. All authors critically reviewed and approved the final version.

## Acknowledgements

This study was supported, in whole or in part, by the Bill & Melinda Gates Foundation (BMGF) and the Coalition for Epidemic Preparedness Innovations (CEPI). The authors thank David P. Figgitt PhD, ISMPP CMPP™, and Katherine F. Croom, Content Ed Net, for providing editorial support during the preparation of the manuscript, with funding from SK Bioscience.

## Supplementary materials

Supplementary material associated with this article can be found in the online version at *medrxiv.org*

## Supplementary Appendix

Supplement to: Cheong, et al. Immunogenicity and safety of SARS-CoV-2 recombinant protein nanoparticle vaccine GBP510 adjuvanted with AS03: randomised, active-controlled, observer-blinded, phase 3 trial

### I Supplementary Tables

**Table S1.** Adverse Events of Special Interest (AESI) Relevant to COVID-19

|  |
| --- |
| **AESI included because they are seen with COVID-19 Disease <sup>3,4</sup>** |
| Acute respiratory distress syndrome |
| Multisystem inflammatory syndrome (children & adults) |
| Acute cardiovascular injury (includes: myocarditis/pericarditis, microangiopathy, heart failure, stress cardiomyopathy, coronary artery disease arrhythmia) |
| Coagulation disorder (includes: thrombotic disorders, bleeding disorders) |
| Anosmia, ageusia |
| Chilblain – like lesions |
| Erythema multiforme |
| Single Organ Cutaneous Vasculitis |
| Acute kidney injury |
| Acute liver injury |
| Acute pancreatitis |
| Rhabdomyolysis |
| Subacute thyroiditis |
| **AESI included because they have a proven or theoretical association with immunization in general** |
| Anaphylaxis <sup>1,2</sup> |
| Thrombocytopenia <sup>1,2,3,4</sup> |
| Generalized convulsion <sup>1,2</sup> |
| Acute disseminated encephalomyelitis <sup>4</sup> |
| Guillain Barré Syndrome <sup>3,4</sup> |
| **AESI included because they have a proven or theoretical association with specific vaccine platform(s)** |
| Acute aseptic arthritis <sup>r-VSV</sup> |
| Aseptic meningitis <sup>Live vaccines</sup> |
| Encephalitis / Encephalomyelitis <sup>Live vaccines</sup> |
| Idiopathic Peripheral Facial Nerve Palsy <sup>Intranasal E. coli Heat Labile Toxin Adjuvanted Vaccine</sup> |
| Vaccine associated enhanced disease <sup>1(Formalin inactivated measles/RSV; HIV), 2(Chimeric YF Dengue), 5 (SARS/MERS-CoVs)</sup> |
| Thrombosis with thrombocytopenia syndrome <sup>Recombinant adenoviral vector vaccines</sup> |

<sup>1</sup> Proven association with immunization encompassing several different vaccines
<sup>2</sup> Proven association with vaccine that could theoretically be true for novel COVID-19 vaccines
<sup>3</sup> Theoretical concern based on wild type disease immunopathogenesis
<sup>4</sup> Theoretical concern related to viral replication during wild type disease
<sup>5</sup> Theoretical concern because it has been demonstrated in an animal model with $\geq 1$ vaccine platform

**Table S2.**
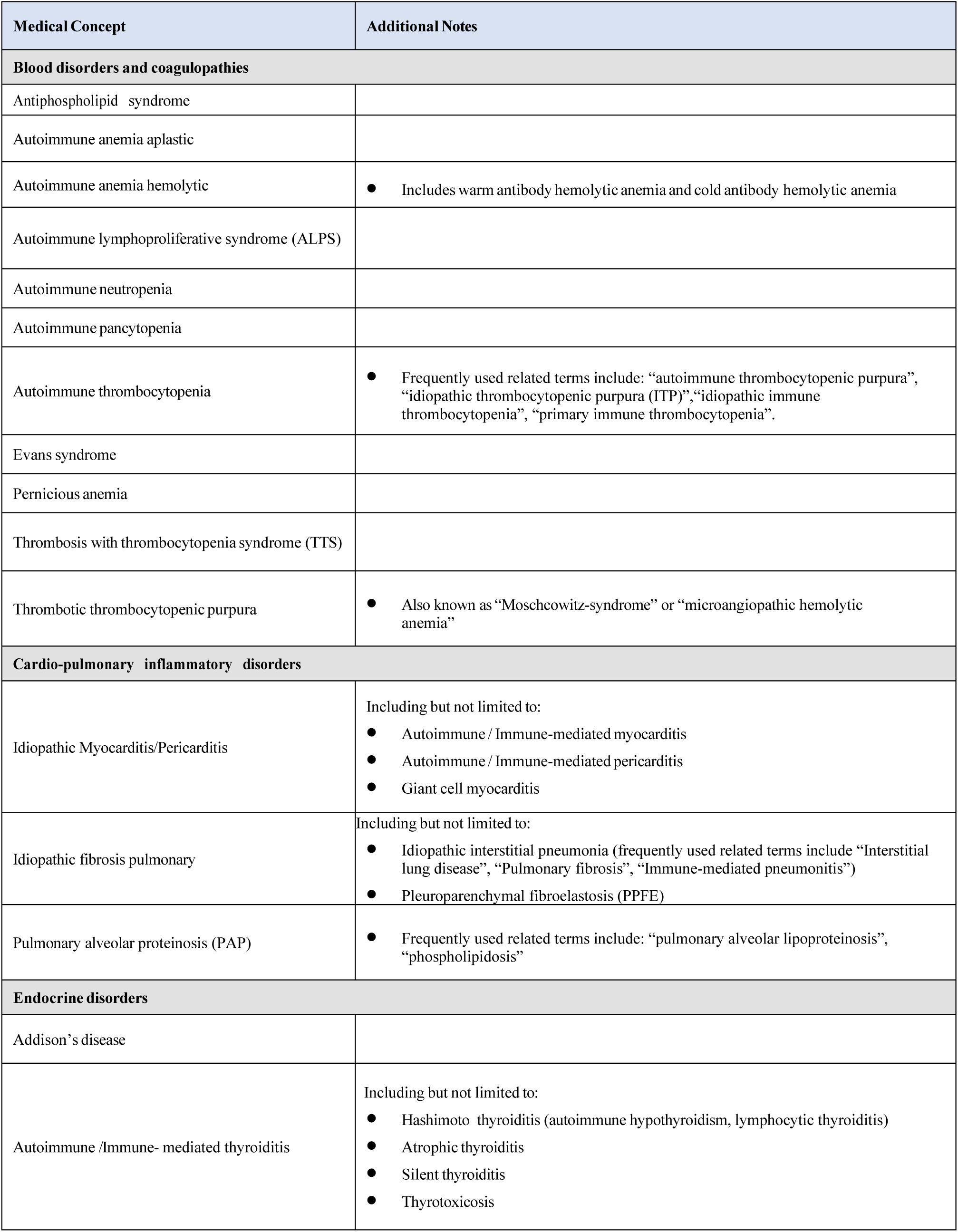
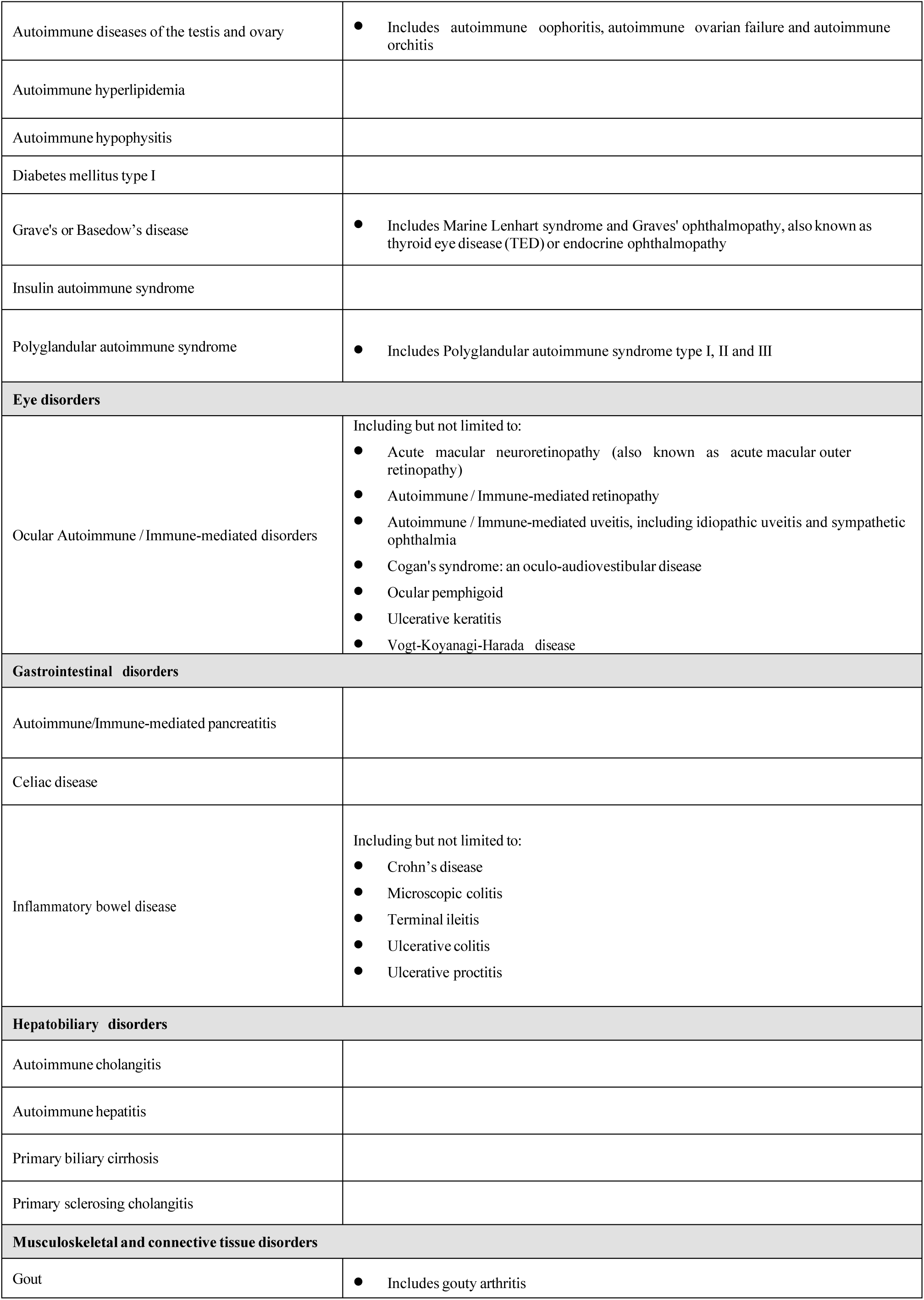
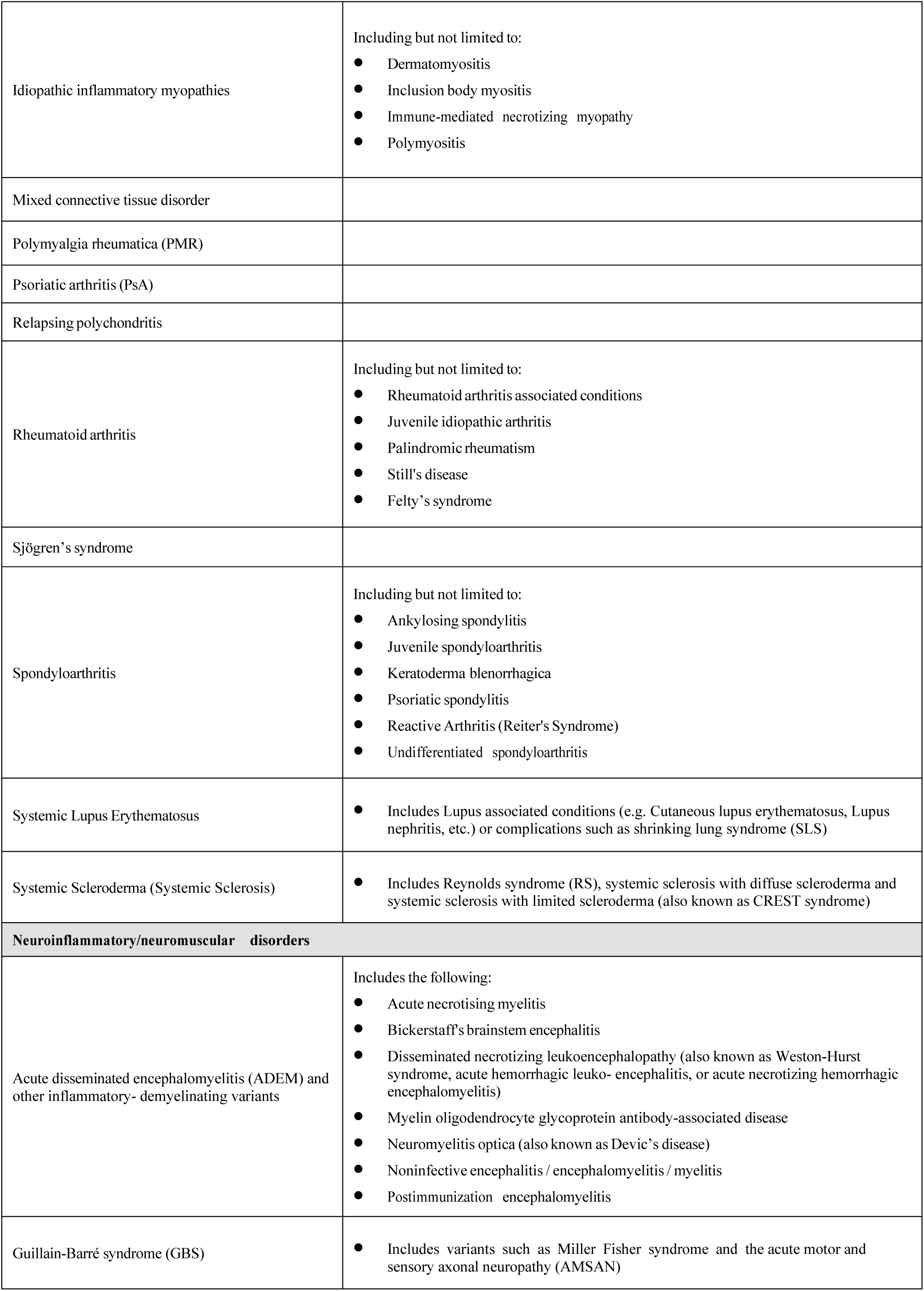
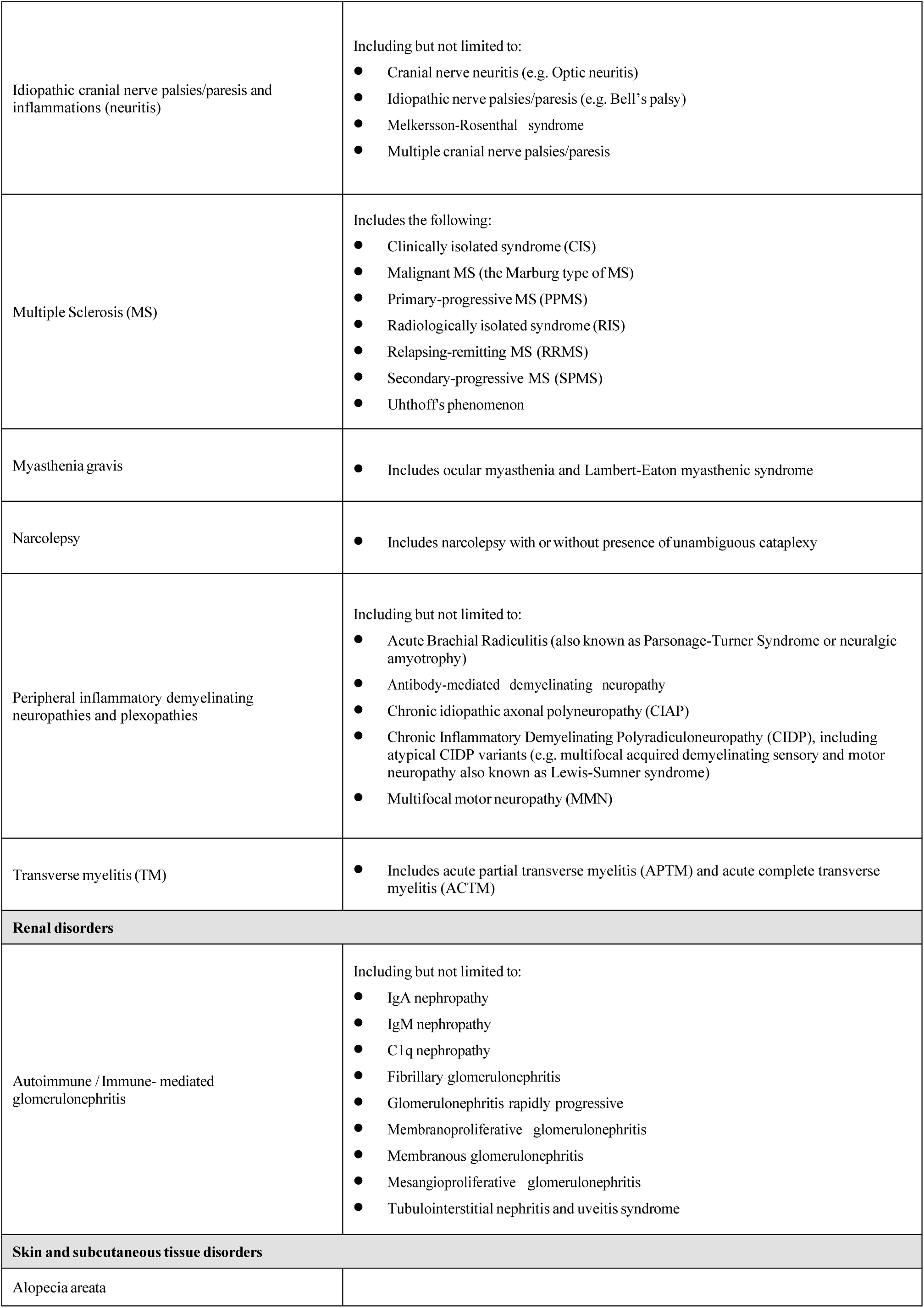
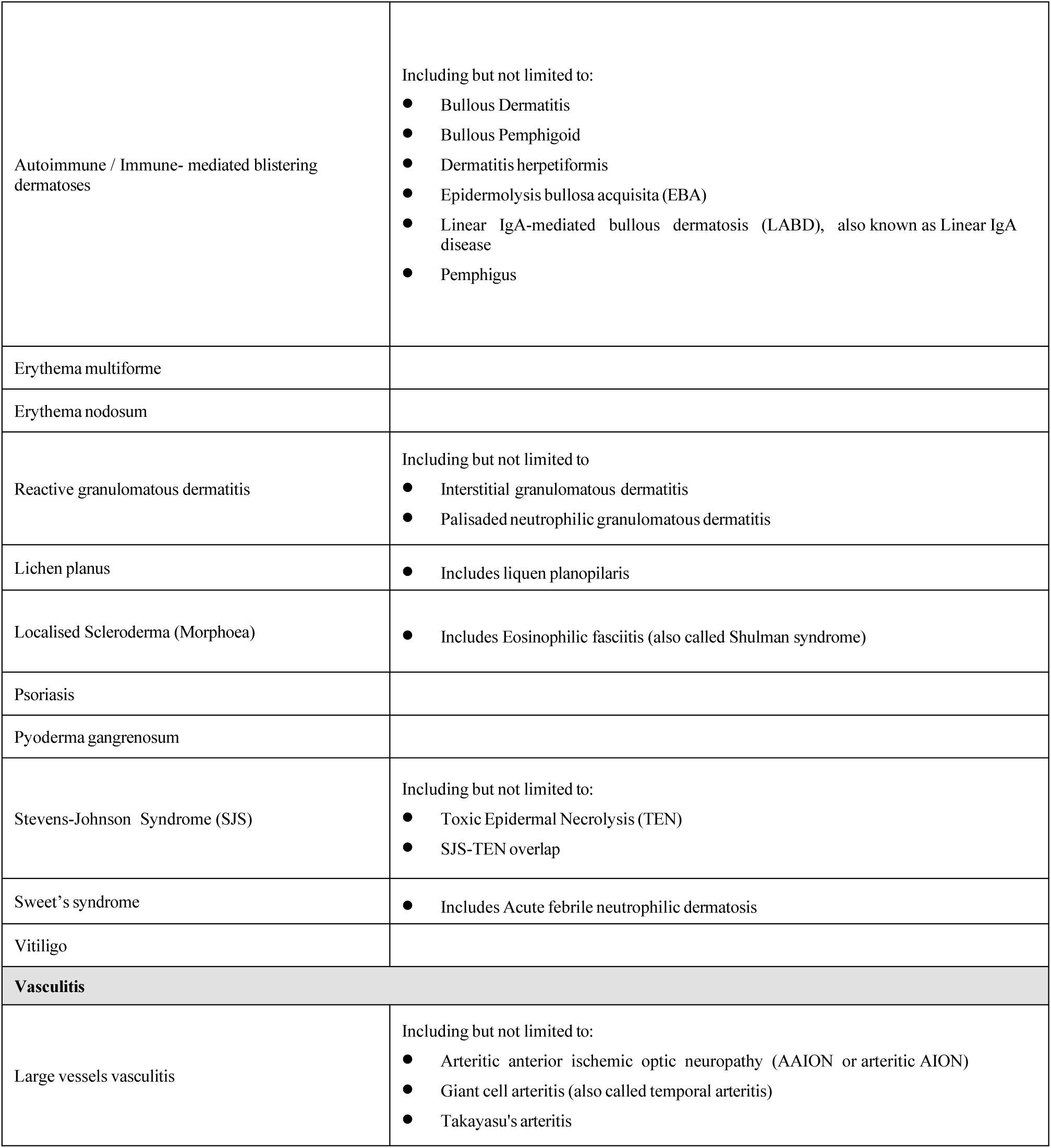
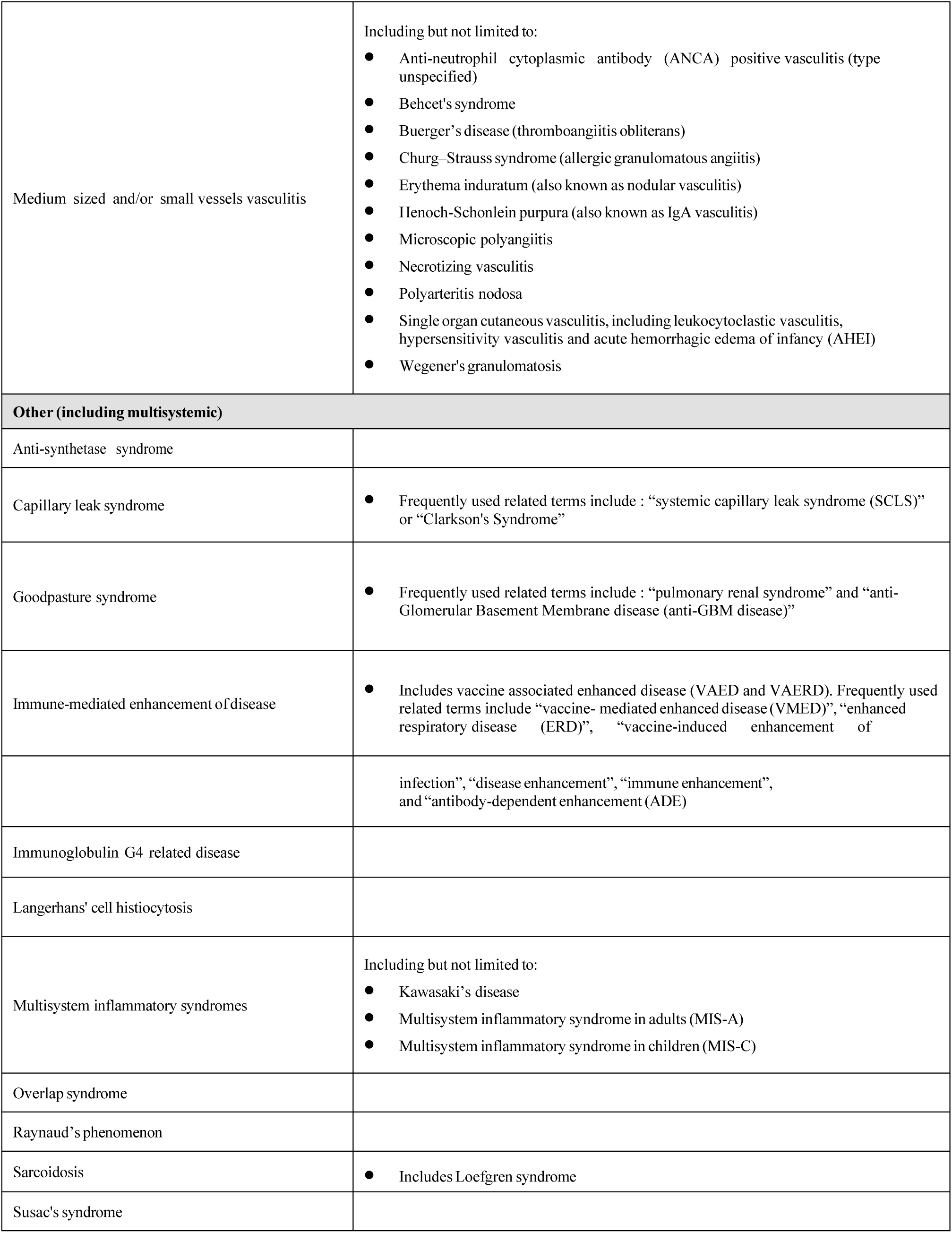
Potential Immune-Mediated Diseases Included as Adverse Events of Special Interest

**Table S3.** Immunogenicity Assessment by Live Virus Neutralisation Assays (FRNT): Age Group 18-64 Years (Per-Protocol Set)

|  | **GBP510**<br>**(N=834)** | **ChAdOx1-S**<br>**(N=417)** |
| --- | --- | --- |
| **Baseline** |  |  |
| n | 834 | 417 |
| GMT(SD) | 8.17(1.08) | 8.13(1.06) |
| 95% Confidence Interval | [8.13, 8.22] | [8.09, 8.18] |
| Ratio of GMTs (GBP510 / ChAdOx1-S)(SD) | 1.01(NA) |  |
| 95% Confidence Interval | [1.00, 1.01] |  |
| P-value [1] | 0.1720 |  |
| **Visit 4 (4 weeks after 1st vaccination)** |  |  |
| n | 187 | 88 |
| GMT(SD) | 11.49(2.28) | 40.74(3.56) |
| 95% Confidence Interval | [10.20, 12.94] | [31.14, 53.30] |
| Ratio of GMTs (GBP510 / ChAdOx1-S)(SD) | 0.28(NA) |  |
| 95% Confidence Interval | [0.21, 0.38] |  |
| P-value [1] | <0.0001 |  |
| GMFR(SD) | 1.41(2.28) | 5.02(3.51) |
| 95% Confidence Interval | [1.26, 1.59] | [3.85, 6.54] |
| Adjusted GMT(SE)† | 11.48(1.07) | 40.79(1.11) |
| 95% Confidence Interval | [9.97, 13.22] | [33.22, 50.10] |
| Ratio of GMTs (GBP510 / ChAdOx1-S)(SE) | 0.28(1.13) |  |
| 95% Confidence Interval | [0.22, 0.36] |  |
| P-value [2] | <0.0001 |  |
| Adjusted GMT(SE)‡ | 9.98(1.15) | 35.51(1.18) |
| 95% Confidence Interval | [7.55, 13.20] | [25.82, 48.82] |
| Ratio of GMTs (GBP510 / ChAdOx1-S)(SE) | 0.28(1.14) |  |
| 95% Confidence Interval | [0.22, 0.36] |  |
| P-value [3] | <0.0001 |  |
| Participants with ≥ 4-fold rise, n(%) | 11(5.88) | 49(55.68) |
| 95% Confidence Interval | [2.97, 10.28] | [44.70, 66.27] |
| Difference in Proportions of the Participant with ≥ 4-fold rise | -49.80 |  |
| 95% Confidence Interval | [-60.85, -38.58] |  |
| P-value [4] | <0.0001 (c) |  |
| **Visit 6 (2 weeks after 2nd vaccination)** |  |  |
| n | 834 | 417 |
| GMT(SD) | 304.52(2.48) | 103.01(2.72) |
| 95% Confidence Interval | [286.29, 323.92] | [93.56, 113.40] |
| Ratio of GMTs (GBP510 / ChAdOx1-S)(SD) | 2.96(NA) |  |
| 95% Confidence Interval | [2.64, 3.31] |  |
| P-value [1] | <0.0001 |  |
| GMFR(SD) | 37.26(2.48) | 12.67(2.72) |
| 95% Confidence Interval | [35.03, 39.63] | [11.51, 13.95] |
| Adjusted GMT(SE)† | 304.31(1.03) | 103.15(1.05) |
| 95% Confidence Interval | [285.49, 324.37] | [94.24, 112.90] |
| Ratio of GMTs (GBP510 / ChAdOx1-S)(SE) | 2.95(1.06) |  |
| 95% Confidence Interval | [2.64, 3.30] |  |
| P-value [2] | <0.0001 |  |
| Adjusted GMT(SE)‡ | 248.59(1.07) | 83.82(1.08) |
| 95% Confidence Interval | [218.85, 282.37] | [72.68, 96.67] |
| Ratio of GMTs (GBP510 / ChAdOx1-S)(SE) | 2.97(1.06) |  |
| 95% Confidence Interval | [2.66, 3.31] |  |
| P-value [3] | <0.0001 |  |
| Participants with $\geq 4$-fold rise, n(%) | 819(98.20) | 366(87.77) |
| 95% Confidence Interval | [97.05, 98.99] | [84.23, 90.76] |
| Difference in Proportions of the Participant with $\geq 4$-fold rise | 10.43 | |
| 95% Confidence Interval | [7.35, 14.06] |  |
| P-value [4] | <0.0001 (c) |  |

GMT = geometric mean titer, SD = standard deviation, SE = standard error, GMFR = geometric mean fold rise, ANCOVA = analysis of covariance, NA = not applicable.
Assay unit: IU/mL
[1] Testing for difference between treatment groups (two sample t-test).
[2] † ANCOVA model with treatment group as a factor, and baseline antibody level as covariate.
[3] ‡ ANCOVA model with treatment group, sex, race as factors and baseline antibody level as covariate.
[4] Testing for difference between treatment groups (chi-square test (c) or Fisher's exact test (f)).
GMT = anti-logarithm[mean of natural logarithm(titer at visit n)], GMFR = anti-logarithm[mean of natural logarithm(titer at visit n/titer at visit 2)].
The test result was collected for randomly selected participants (about 20% of all participants) at Visit 4.
When two samples had unequal variance, the standard deviation for the ratio of GMT is presented 'NA'.
Due to infinite likelihood problem, the variance model for adjusted GMT was fitted under the assumption of homoscedasticity.

**Table S4.**
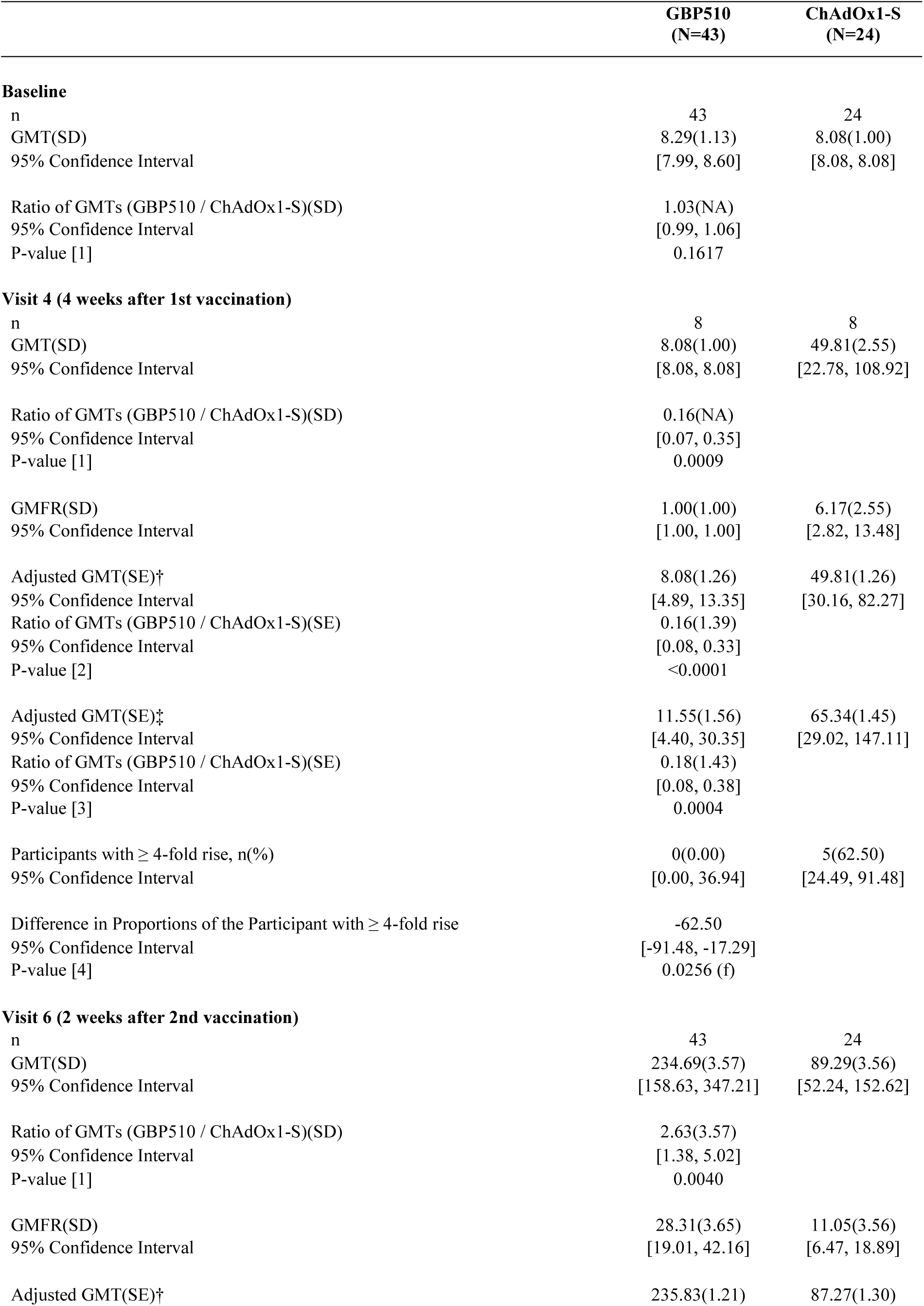
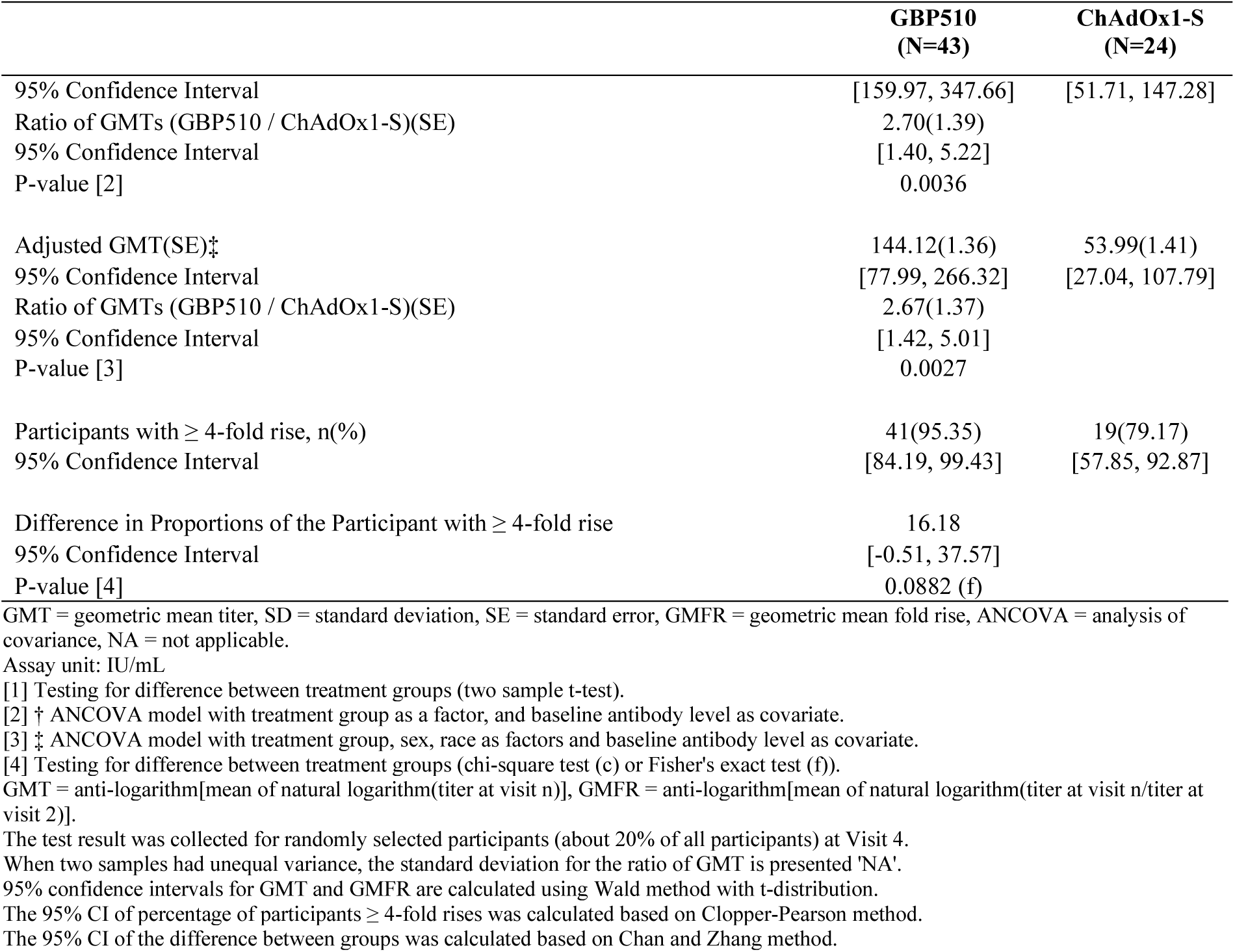
Immunogenicity Assessment by Live Virus Neutralisation Assays (FRNT): Age Group ≥65 Years (Per-Protocol Set)

**Table S5.** The Consistency Among Ethnicities by Live Virus Neutralisation Assay (FRNT) (Per Protocol Set)

|  | Korean |  | Southeast Asian |  | Caucasian |  |
| --- | --- | --- | --- | --- | --- | --- |
|  | GBP510<br>(N=303) | ChAdOx1-S<br>(N=154) | GBP510<br>(N=529) | ChAdOx1-S<br>(N=266) | GBP510<br>(N=45) | ChAdOx1-S<br>(N=21) |
| **Baseline** |  |  |  |  |  |  |
| n | 303 | 154 | 529 | 266 | 45 | 21 |
| GMT(SD) | 8.20(1.09) | 8.14(1.07) | 8.16(1.08) | 8.12(1.05) | 8.24(1.10) | 8.08(1.00) |
| 95% Confidence Interval | [8.12, 8.28] | [8.06, 8.23] | [8.11, 8.21] | [8.07, 8.17] | [8.01, 8.47] | [8.08, 8.08] |
| Ratio of GMTs (GBP510 / ChAdOx1-S)(SD) | 1.01(NA) |  | 1.00(NA) |  | 1.02(NA) |  |
| 95% Confidence Interval | [0.99, 1.02] |  | [1.00, 1.01] |  | [0.99, 1.05] |  |
| P-value [1] | 0.3169 |  | 0.2992 |  | 0.1597 |  |
| **Visit 6 (2 weeks after 2nd vaccination)** |  |  |  |  |  |  |
| n | 303 | 154 | 529 | 266 | 45 | 21 |
| GMT(SD) | 306.47(2.17) | 75.26(2.54) | 305.79(2.68) | 131.12(2.67) | 216.59(3.23) | 41.12(2.48) |
| 95% Confidence Interval | [280.82, 334.47] | [64.89, 87.29] | [281.12, 332.62] | [116.48, 147.60] | [152.29, 308.04] | [27.17, 62.21] |
| Ratio of GMTs (GBP510 / ChAdOx1-S)(SD) | 4.07(NA) |  | 2.33(2.67) |  | 5.27(3.00) |  |
| 95% Confidence Interval | [3.43, 4.84] |  | [2.02, 2.70] |  | [2.95, 9.40] |  |
| P-value [1] | <0.0001 |  | <0.0001 |  | <0.0001 |  |
| GMFR(SD) | 37.38(2.17) | 9.25(2.54) | 37.47(2.68) | 16.14(2.67) | 26.28(3.31) | 5.09(2.48) |
| 95% Confidence Interval | [34.24, 40.80] | [7.97, 10.72] | [34.44, 40.76] | [14.34, 18.17] | [18.34, 37.67] | [3.36, 7.70] |
| Adjusted GMT(SE)† | 528.33(1.27) | 129.88(1.28) | 268.71(1.07) | 115.88(1.09) | 154.40(1.62) | 28.11(1.65) |
| 95% Confidence Interval | [328.71, 849.20] | [80.10, 210.62] | [233.68, 308.99] | [98.64, 136.12] | [59.19, 402.80] | [10.32, 76.60] |
| Ratio of GMTs (GBP510 / ChAdOx1-S)(SE) | 4.07(1.08) |  | 2.32(1.08) |  | 5.49(1.31) |  |
| 95% Confidence Interval | [3.47, 4.77] |  | [2.01, 2.68] |  | [3.21, 9.38] |  |
| P-value [2] | <0.0001 |  | <0.0001 |  | <0.0001 |  |
| Ratio of GMTs > 1 | Yes |  | Yes |  | Yes |  |
| Adjusted GMT(SE)‡ | 522.57(1.28) | 128.49(1.28) | 270.12(1.07) | 115.96(1.09) | 144.68(1.36) | 27.77(1.41) |
| 95% Confidence Interval | [324.01, 842.80] | [78.97, 209.07] | [234.87, 310.65] | [98.72, 136.21] | [77.66, 269.53] | [14.06, 54.83] |
| Ratio of GMTs (GBP510 / ChAdOx1-S)(SE) | 4.07(1.09) |  | 2.33(1.08) |  | 5.21(1.31) |  |
| 95% Confidence Interval | [3.46, 4.77] |  | [2.02, 2.69] |  | [3.05, 8.91] |  |
| P-value [3] | <0.0001 |  | <0.0001 |  | <0.0001 |  |
|  | GBP510<br>(N=303) | ChAdOx1-S<br>(N=154) | GBP510<br>(N=529) | ChAdOx1-S<br>(N=266) | GBP510<br>(N=45) | ChAdOx1-S<br>(N=21) |
| Participants with $\geq 4$-fold rise, n(%) | 301(99.34) | 124(80.52) | 518(97.92) | 249(93.61) | 41(91.11) | 12(57.14) |
| 95% Confidence Interval | [97.64, 99.92] | [73.37, 86.45] | [96.31, 98.96] | [89.96, 96.23] | [78.78, 97.52] | [34.02, 78.18] |
| Difference in Proportions of the Participant with $\geq 4$-fold rise | 18.82 | | 4.31 | | 33.97 | |
| 95% Confidence Interval | [12.50, 25.14] |  | [1.13, 7.49] |  | [11.23, 56.71] |  |
| P-value [4] | <0.0001 (c) |  | 0.0019 (c) |  | 0.0024 (f) |  |
| Difference in Proportions of the Participant with $\geq 4$-fold rise > -5% | Yes | | Yes | | Yes | |

GMT = geometric mean titer, SD = standard deviation, SE = standard error, GMFR = geometric mean fold rise, ANCOVA = analysis of covariance, NA = not applicable.
Assay unit: IU/mL
[1] Testing for between-treatment groups (two sample t-test).
[2] † ANCOVA model with treatment group, age group (18~64, $\geq 65$ ) as factors, and baseline antibody level as covariate.
[3] ‡ ANCOVA model with treatment group, age group (18~64, $\geq 65$ ), sex as factors and baseline antibody level as covariate.
[4] Testing for difference between treatment groups (chi-square test (c) or Fisher's exact test (f)).
GMT = anti-logarithm[mean of natural logarithm(titer at Visit n)], GMFR = anti-logarithm[mean of natural logarithm(titer at visit n/titer at visit 2)].
When two samples had unequal variance, the standard deviation for the ratio of GMT is presented 'NA'.
95% confidence interval for GMT and GMFR are calculated using t-distribution and Confidence Interval of Seroconversion rate is calculated using Wald method.
95% confidence interval for the percentage of participants $\geq 4$ -fold rises was calculated by Clopper-Pearson Methods.

**Table S6.** Immunogenicity Assessment by Live Virus Neutralization Assays (FRNT50) against Delta (Per-Protocol Set 1)

|  | **GBP510**<br>**(N=877)** | **ChAdOx1-S**<br>**(N=441)** |
| --- | --- | --- |
| **Visit 6 (2 weeks after 2nd vaccination)** |  |  |
| n | 137 | 68 |
| GMT(SD) | 2644.26(2.47) | 96.96(4.01) |
| 95% Confidence Interval | [2269.54, 3080.84] | [69.29, 135.67] |
| Ratio of GMTs (GBP510 / ChAdOx1-S)(SD) | 27.27(NA) |  |
| 95% Confidence Interval | [18.88, 39.39] |  |
| P-value [1] | <0.0001 |  |

GMT = geometric mean titer, SD = standard deviation, NA = not applicable.
GMT = anti-logarithm[mean of natural logarithm(titer at visit n)].
The test result was collected for randomly selected participants (about 10% of all participants).
[1] Testing for between-treatment groups (two sample t-test).
When two samples had unequal variance, the standard deviation for the ratio of GMT is presented 'NA'.
95% confidence intervals for GMT are calculated using Wald method with t-distribution.

**Table S7.**
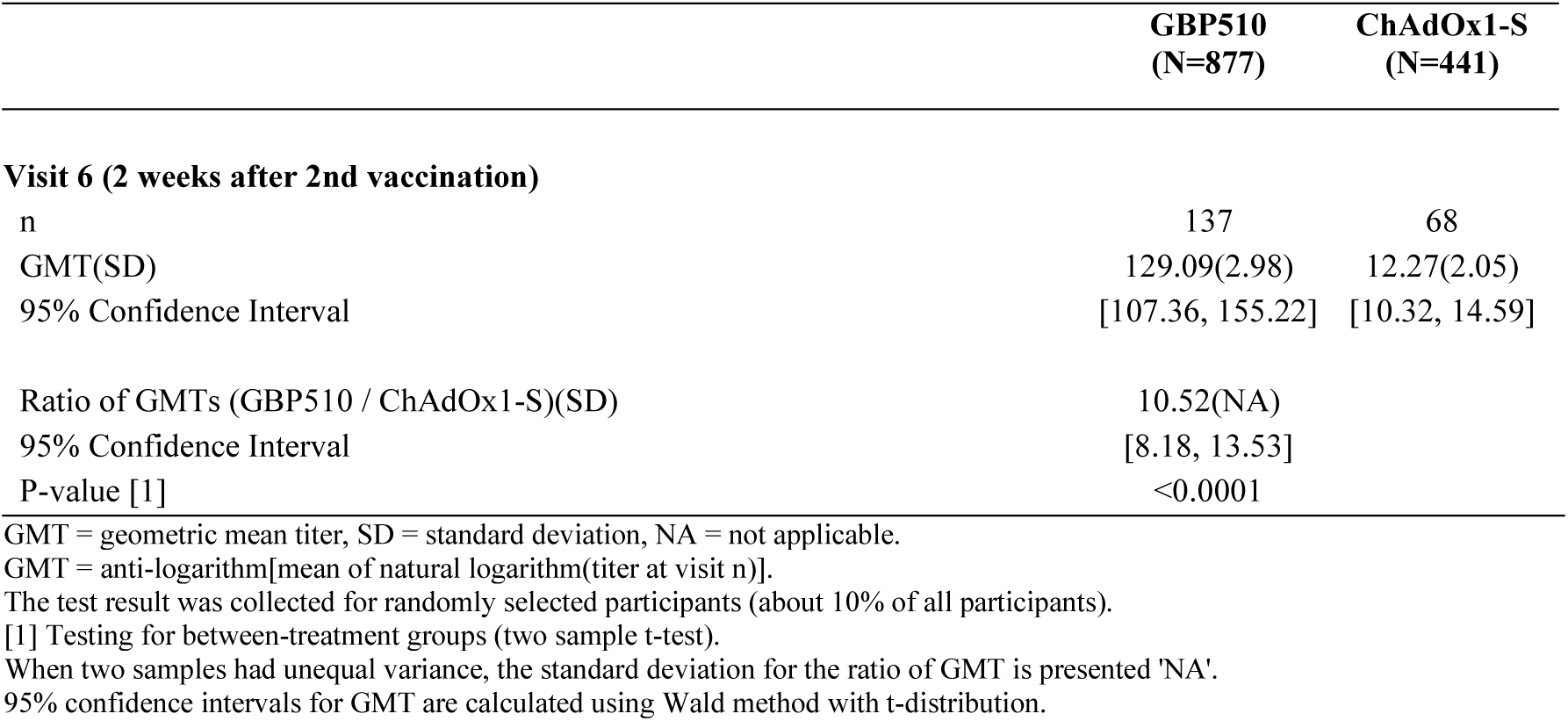
Immunogenicity Assessment by Live Virus Neutralizing Assays (FRNT50) against Omicron BA.1 (Per-Protocol Set 1)

**Table S8.** Immunogenicity Assessment by Live Virus Neutralizing Assays (FRNT50) against Omicron BA. 5 (Per-Protocol Set 1)

|  | **GBP510**<br>**(N=877)** | **ChAdOx1-S**<br>**(N=441)** |
| --- | --- | --- |
| **Visit 6 (2 weeks after 2nd vaccination)** |  |  |
| n | 137 | 68 |
| GMT(SD) | 61.61(2.84) | 13.56(2.19) |
| 95% Confidence Interval | [51.64, 73.51] | [11.21, 16.40] |
| Ratio of GMTs (GBP510 / ChAdOx1-S)(SD) | 4.54(NA) |  |
| 95% Confidence Interval | [3.51, 5.88] |  |
| P-value [1] | <0.0001 |  |

GMT = geometric mean titer, SD = standard deviation, NA = not applicable.
GMT = anti-logarithm[mean of natural logarithm(titer at visit n)].
The test result was collected for randomly selected participants (about 10% of all participants).
[1] Testing for between-treatment groups (two sample t-test).
When two samples had unequal variance, the standard deviation for the ratio of GMT is presented 'NA'.
95% confidence intervals for GMT are calculated using Wald method with t-distribution.

**Table S9.** Cell-mediated Response: CD8+ T cell at Visit 6 (Per-Protocol Set 1)

|  | **GBP510**<br>**(N=877)** | **ChAdOx1-S**<br>**(N=441)** |
| --- | --- | --- |
| **[cytokines: IFNγ]** |  |  |
| **Baseline** |  |  |
| n | 70 | 36 |
| Mean(SD) | 0.00(0.01) | 0.00(0.00) |
| Median | 0.00 | 0.00 |
| Min, Max | 0.00, 0.05 | 0.00, 0.02 |
| P-value [1] | 0.3052 |  |
| n | 24 | 10 |
| Geometric Mean(SD) | 0.01(1.86) | 0.01(1.60) |
| 95% Confidence Interval | [0.01, 0.01] | [0.01, 0.01] |
| P-value [1] | 0.7495 |  |
| **Visit 6 (2 weeks after 2nd vaccination)** |  |  |
| n | 68 | 33 |
| Mean(SD) | 0.01(0.02) | 0.01(0.01) |
| Median | 0.00 | 0.01 |
| Min, Max | 0.00, 0.15 | 0.00, 0.06 |
| P-value [1] | 0.8398 |  |
| n | 22 | 18 |
| Geometric Mean(SD) | 0.01(2.79) | 0.01(1.96) |
| 95% Confidence Interval | [0.01, 0.02] | [0.01, 0.01] |
| P-value [1] | 0.3523 |  |
| **[cytokines: IL-2]** |  |  |
| **Baseline** |  |  |
| n | 70 | 36 |
| Mean(SD) | 0.00(0.01) | 0.00(0.01) |
| Median | 0.00 | 0.00 |
| Min, Max | 0.00, 0.05 | 0.00, 0.06 |
| P-value [1] | 0.6094 |  |
| n | 25 | 7 |
| Geometric Mean(SD) | 0.01(2.01) | 0.01(2.71) |
| 95% Confidence Interval | [0.01, 0.01] | [0.00, 0.02] |
| P-value [1] | 0.8245 |  |
| **Visit 6 (2 weeks after 2nd vaccination)** |  |  |
| n | 68 | 33 |
| Mean(SD) | 0.00(0.01) | 0.01(0.01) |
| Median | 0.00 | 0.00 |
| Min, Max | 0.00, 0.08 | 0.00, 0.03 |
| P-value [1] | 0.4842 |  |
| n | 20 | 15 |
| Geometric Mean(SD) | 0.01(2.05) | 0.01(2.03) |
| 95% Confidence Interval | [0.01, 0.02] | [0.01, 0.02] |
| P-value [1] | 0.7884 |  |
| **[cytokines: IL-4]** |  |  |
| **Baseline** |  |  |
| n | 70 | 36 |
| Mean(SD) | 0.01(0.01) | 0.01(0.01) |
| Median | 0.00 | 0.00 |
| Min, Max | 0.00, 0.07 | 0.00, 0.04 |
| P-value [1] | 0.9768 |  |
| n | 24 | 11 |
| Geometric Mean(SD) | 0.01(2.12) | 0.01(2.11) |
| 95% Confidence Interval | [0.01, 0.01] | [0.01, 0.02] |
| P-value [1] | 0.4107 |  |
| **Visit 6 (2 weeks after 2nd vaccination)** |  |  |
| n | 68 | 33 |
| Mean(SD) | 0.01(0.01) | 0.00(0.01) |
| Median | 0.00 | 0.00 |
| Min, Max | 0.00, 0.06 | 0.00, 0.03 |
| P-value [1] | 0.4652 |  |
| n | 29 | 11 |
| Geometric Mean(SD) | 0.01(1.81) | 0.01(1.92) |
| 95% Confidence Interval | [0.01, 0.01] | [0.01, 0.02] |
| P-value [1] | 0.8626 |  |
| **[cytokines: TNFα]** |  |  |
| **Baseline** |  |  |
| n | 70 | 36 |
| Mean(SD) | 0.04(0.12) | 0.04(0.12) |
| Median | 0.02 | 0.02 |
| Min, Max | 0.00, 0.88 | 0.00, 0.73 |
| P-value [1] | 0.9816 |  |
| n | 45 | 22 |
| Geometric Mean(SD) | 0.03(3.14) | 0.03(2.77) |
| 95% Confidence Interval | [0.02, 0.04] | [0.02, 0.05] |
| P-value [1] | 0.6388 |  |
| **Visit 6 (2 weeks after 2nd vaccination)** |  |  |
| n | 68 | 33 |
| Mean(SD) | 0.11(0.26) | 0.07(0.13) |
| Median | 0.02 | 0.02 |
| Min, Max | 0.00, 1.66 | 0.00, 0.57 |
| P-value [1] | 0.2742 |  |
| n | 41 | 24 |
| Geometric Mean(SD) | 0.08(3.81) | 0.04(4.34) |
| 95% Confidence Interval | [0.05, 0.12] | [0.02, 0.07] |
| P-value [1] | 0.0419 |  |

SD = standard deviation, Min = minimum, Max = maximum, CMI = cell mediated immunity.
Assay unit: % of RBD-specific CD8+ T cells
[1] Testing for difference between treatment groups (two sample t-test).
The test result was collected for randomly selected participants (about 10% of all participants).
Geometric Mean = anti-logarithm[mean of natural logarithm(percentage of RBD-specific CD8+ T-cell at visit n)].
The 95% confidence intervals for Geometric Mean are calculated using Wald method with t-distribution.

**Table S10.** Solicited Local AE by Maximum severity [after any vaccination] (Safety Set)

|  | GBP510<br>(N=3029) |  |  |  | ChAdOx1-S<br>(N=996) |  |  |  |
| --- | --- | --- | --- | --- | --- | --- | --- | --- |
|  | Grade 1<br>(Mild) | Grade 2<br>(Moderate) | Grade 3<br>(Severe) | Grade 4<br>(Potentially Life<br>Threatening) | Grade 1<br>(Mild) | Grade 2<br>(Moderate) | Grade 3<br>(Severe) | Grade 4<br>(Potentially Life<br>Threatening) |
| **Adverse Events after any**<br>**vaccination, n(%)** |  |  |  |  |  |  |  |  |
| Injection Site Pain | 1348(44.50) | 293(9.67) | 46(1.52) | 0 | 414(41.57) | 66(6.63) | 6(0.60) | 0 |
| Injection Site Redness | 101(3.33) | 49(1.62) | 9(0.30) | 0 | 20(2.01) | 0 | 0 | 0 |
| Injection Site Swelling | 123(4.06) | 42(1.39) | 3(0.10) | 0 | 11(1.10) | 1(0.10) | 0 | 0 |

Note: Denominator of percentage is the number of participants in each group.
Adverse events are displayed as 'number of participants(percentage of participants)'.
If one subject experienced the same adverse event more than once, the adverse event is counted only once with the most severe category.

**Table S11.** Solicited Systemic AEs by Maximum Severity [after any vaccination] (Safety Set)

|  | GBP510<br>(N=3029) |  |  |  | ChAdOx1-S<br>(N=996) |  |  |  |
| --- | --- | --- | --- | --- | --- | --- | --- | --- |
|  | Grade 1<br>(Mild) | Grade 2<br>(Moderate) | Grade 3<br>(Severe) | Grade 4<br>(Potentially Life<br>Threatening) | Grade 1<br>(Mild) | Grade 2<br>(Moderate) | Grade 3<br>(Severe) | Grade 4<br>(Potentially Life<br>Threatening) |
| **Adverse Events after any**<br>**vaccination, n(%)** |  |  |  |  |  |  |  |  |
| Fever | 260(8.58) | 159(5.25) | 55(1.82) | 0 | 88(8.84) | 45(4.52) | 28(2.81) | 1(0.10) |
| Nausea/Vomiting | 178(5.88) | 25(0.83) | 0 | 0 | 63(6.33) | 14(1.41) | 1(0.10) | 0 |
| Diarrhea | 136(4.49) | 28(0.92) | 3(0.10) | 0 | 70(7.03) | 10(1.00) | 0 | 0 |
| Headache | 680(22.45) | 200(6.60) | 25(0.83) | 0 | 258(25.90) | 65(6.53) | 8(0.80) | 0 |
| Fatigue | 626(20.67) | 253(8.35) | 62(2.05) | 0 | 205(20.58) | 101(10.14) | 21(2.11) | 0 |
| Myalgia | 553(18.26) | 306(10.10) | 64(2.11) | 0 | 198(19.88) | 94(9.44) | 15(1.51) | 0 |
| Arthralgia | 295(9.74) | 126(4.16) | 26(0.86) | 0 | 120(12.05) | 48(4.82) | 8(0.80) | 0 |
| Chills | 456(15.05) | 180(5.94) | 49(1.62) | 0 | 155(15.56) | 70(7.03) | 21(2.11) | 0 |

Note: Denominator of percentage is the number of participants in each group.
Adverse events are displayed as 'number of participants(percentage of participants)'.
If one subject experienced the same adverse event more than once, the adverse event is counted only once with the most severe category.

**Table S12.**
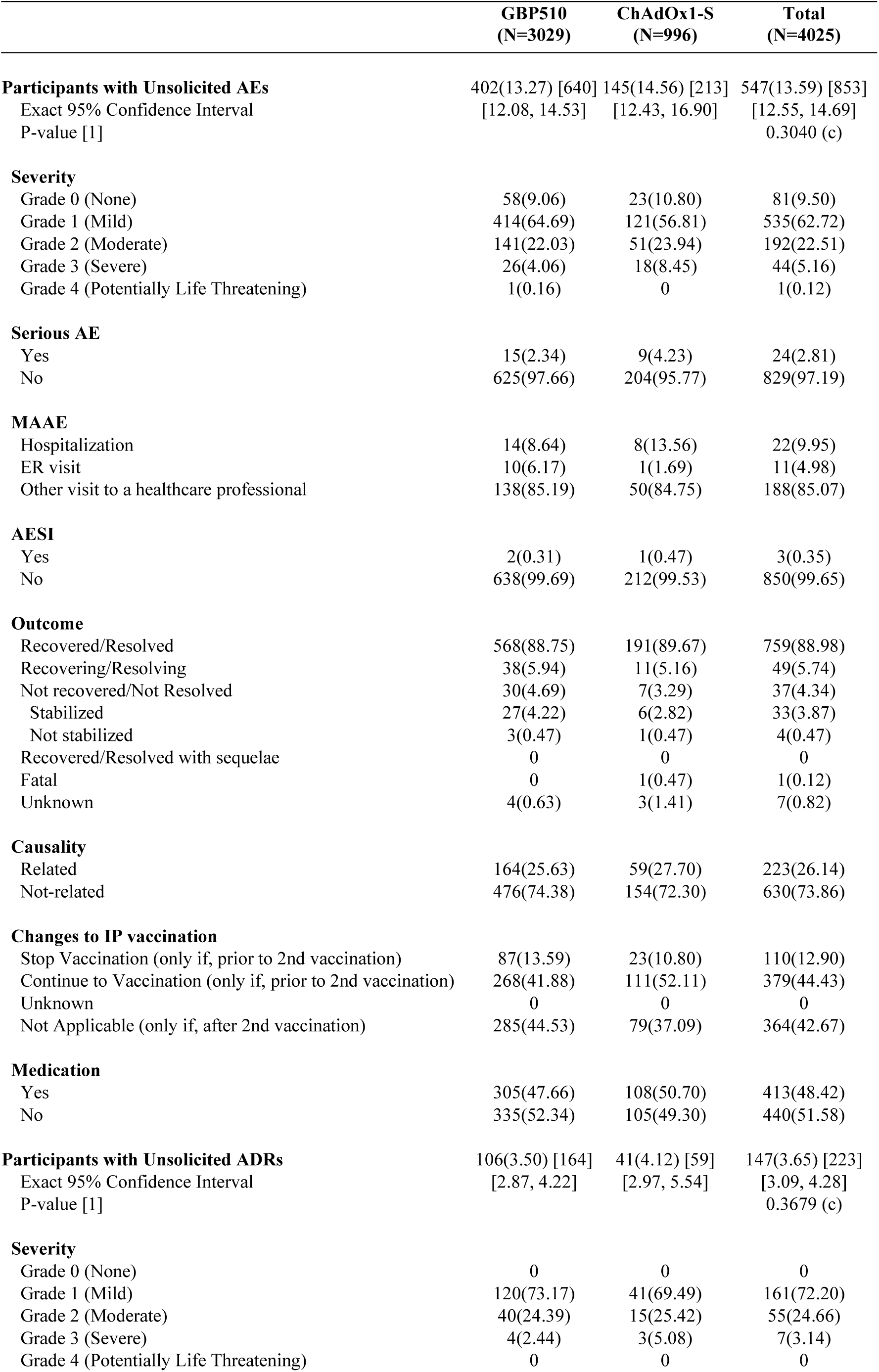
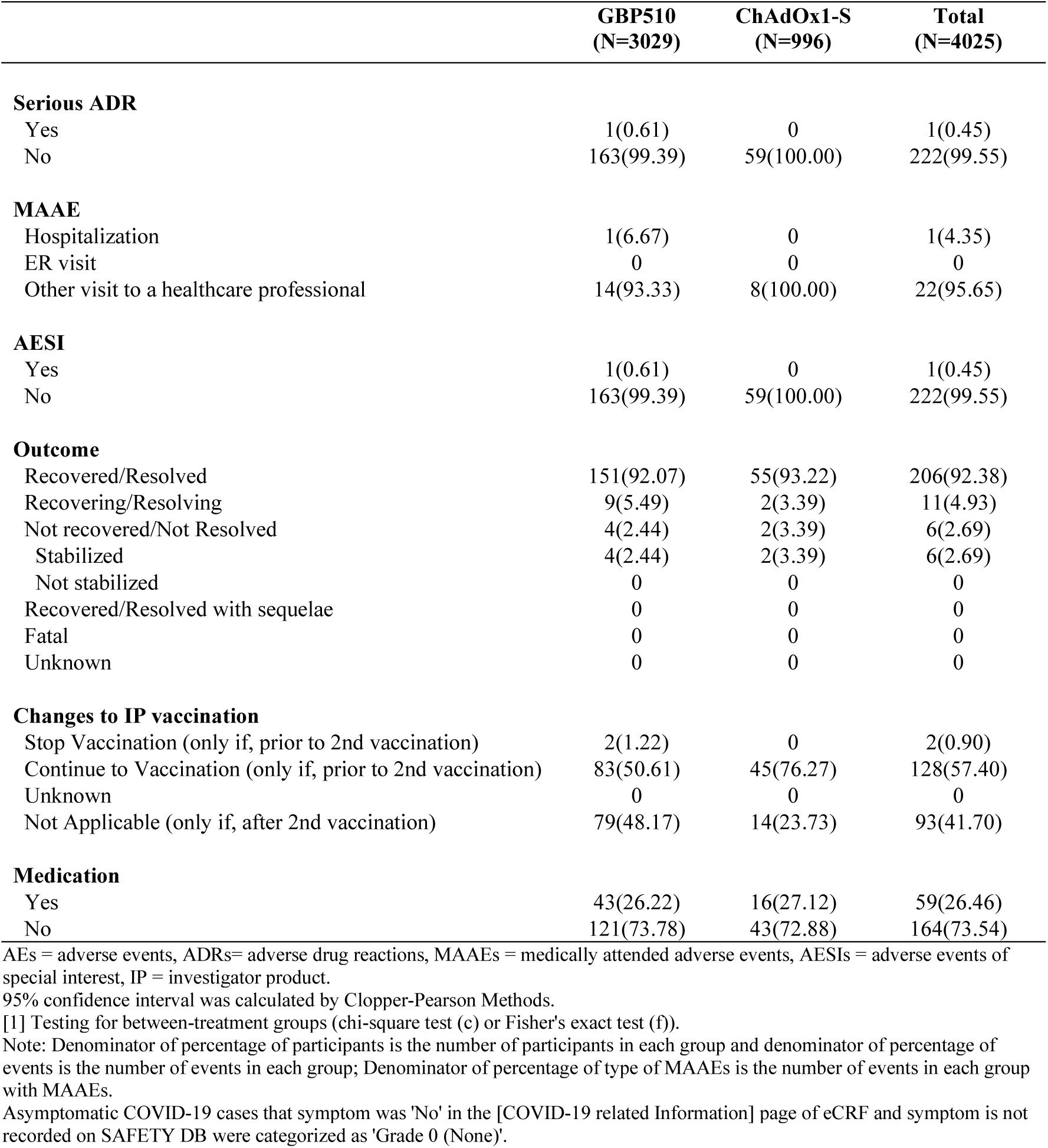
Unsolicited AE [after any vaccination] (Safety Set)

**Table S13.** Virologically-confirmed COVID-19 Observed within a Median 2.5-Months of Follow-up after 2nd Vaccination (Safety Set)

|  | **GBP510**<br>**(N=3029)** | **ChAdOx1-S**<br>**(N=996)** |
| --- | --- | --- |
| **After any vaccination** | 152(5.02) | 77(7.73) |
| Exact 95% Confidence Interval | [4.27, 5.86] | [6.15, 9.57] |
| P-value [1] | 0.0013 (c) |  |
| **Severity** |  |  |
| Asymptomatic | 30(0.99) | 19(1.91) |
| Non-severe | 122(4.03) | 58(5.82) |
| Severe | 0 | 0 |
| Critical | 0 | 0 |
| **<14 days after 1st vaccination** | 9(0.30) | 1(0.10) |
| Exact 95% Confidence Interval | [0.14, 0.56] | [0.00, 0.56] |
| P-value [1] | 0.4674 (f) |  |
| **Severity** |  |  |
| Asymptomatic | 3(0.10) | 0 |
| Non-severe | 6(0.20) | 1(0.10) |
| Severe | 0 | 0 |
| Critical | 0 | 0 |
| **≥ 14 days after 1st vaccination and before 2nd vaccination** | 32(1.06) | 15(1.51) |
| Exact 95% Confidence Interval | [0.72, 1.49] | [0.85, 2.47] |
| P-value [1] | 0.2519 (c) |  |
| **Severity** |  |  |
| Asymptomatic | 10(0.33) | 7(0.70) |
| Non-severe | 22(0.73) | 8(0.80) |
| Severe | 0 | 0 |
| Critical | 0 | 0 |
| **After 2nd vaccination and <14 days after 2nd vaccination** | 6(0.20) | 2(0.20) |
| Exact 95% Confidence Interval | [0.07, 0.43] | [0.02, 0.72] |
| P-value [1] | 1.0000 (f) |  |
| **Severity** |  |  |
| Asymptomatic | 3(0.10) | 0 |
| Non-severe | 3(0.10) | 2(0.20) |
| Severe | 0 | 0 |
| Critical | 0 | 0 |
| **≥ 14 days after 2nd vaccination** | 105(3.47) | 59(5.92) |
| Exact 95% Confidence Interval | [2.84, 4.18] | [4.54, 7.57] |
| P-value [1] | 0.0007 (c) |  |
| **Severity** |  |  |
| Asymptomatic | 14(0.46) | 12(1.20) |
| Non-severe | 91(3.00) | 47(4.72) |
| Severe | 0 | 0 |
| Critical | 0 | 0 |

**Table S14.** Virologically-suspected COVID-19 Observed within a Median 2.5-Months of Follow-up after 2nd Vaccination (Safety set)

|  | **GBP510**<br>**(N=3029)** | **ChAdOx1-S**<br>**(N=996)** |
| --- | --- | --- |
| **After any vaccination** | 86(2.84) | 21(2.11) |
| Exact 95% Confidence Interval | [2.28, 3.49] | [1.31, 3.20] |
| P-value [1] | 0.2136 (c) |  |
| **Severity** |  |  |
| Asymptomatic | 39(1.29) | 7(0.70) |
| Non-severe | 47(1.55) | 14(1.41) |
| Severe | 0 | 0 |
| Critical | 0 | 0 |
| **<14 days after 1st vaccination** | 0 | 0 |
| Exact 95% Confidence Interval | [0.00, 0.12] | [0.00, 0.37] |
| P-value [1] | NC |  |
| **≥ 14 days after 1st vaccination and before 2nd vaccination** | 36(1.19) | 5(0.50) |
| Exact 95% Confidence Interval | [0.83, 1.64] | [0.16, 1.17] |
| P-value [1] | 0.0612 (c) |  |
| **Severity** |  |  |
| Asymptomatic | 33(1.09) | 5(0.50) |
| Non-severe | 3(0.10) | 0 |
| Severe | 0 | 0 |
| Critical | 0 | 0 |
| **After 2nd vaccination and <14 days after 2nd vaccination** | 0 | 0 |
| Exact 95% Confidence Interval | [0.00, 0.12] | [0.00, 0.37] |
| P-value [1] | NC |  |
| **≥ 14 days after 2nd vaccination** | 50(1.65) | 16(1.61) |
| Exact 95% Confidence Interval | [1.23, 2.17] | [0.92, 2.60] |
| P-value [1] | 0.9239 (c) |  |
| **Severity** |  |  |
| Asymptomatic | 6(0.20) | 2(0.20) |
| Non-severe | 44(1.45) | 14(1.41) |
| Severe | 0 | 0 |
| Critical | 0 | 0 |

NC = not calculated.

### II Supplementary Figures

**Figure S1.**
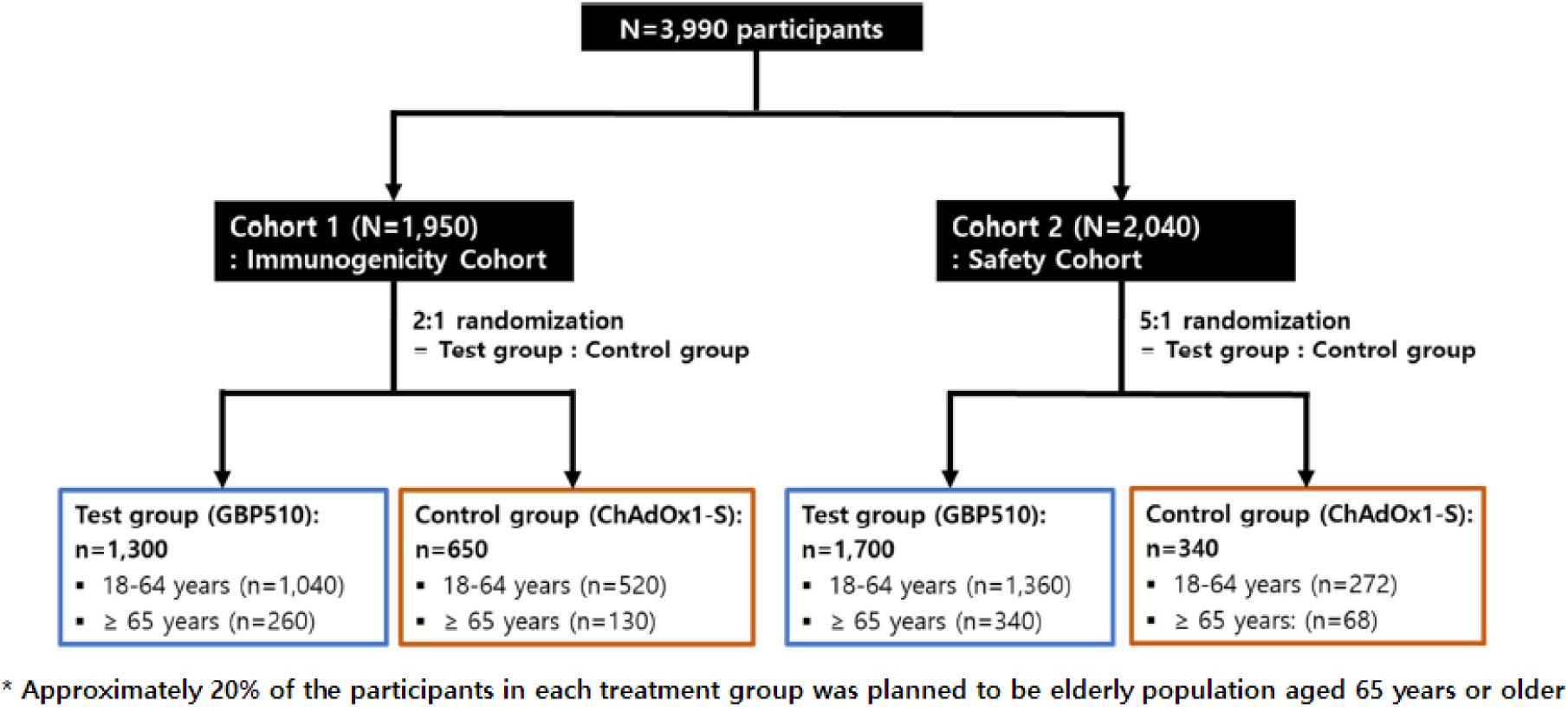
Schema of Study Enrolment

**Figure S2.**
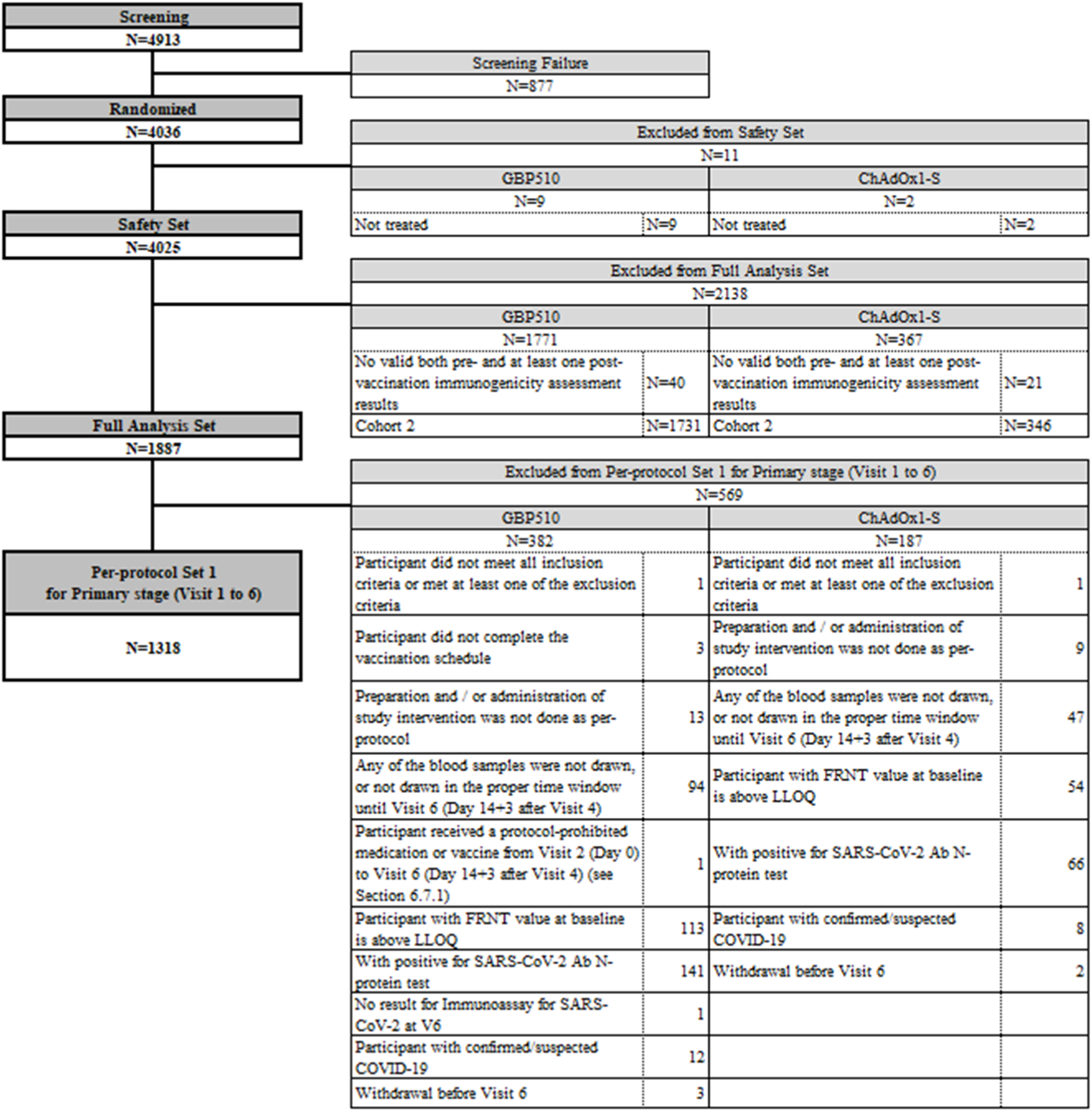
Analysis Sets

**Figure S3.**
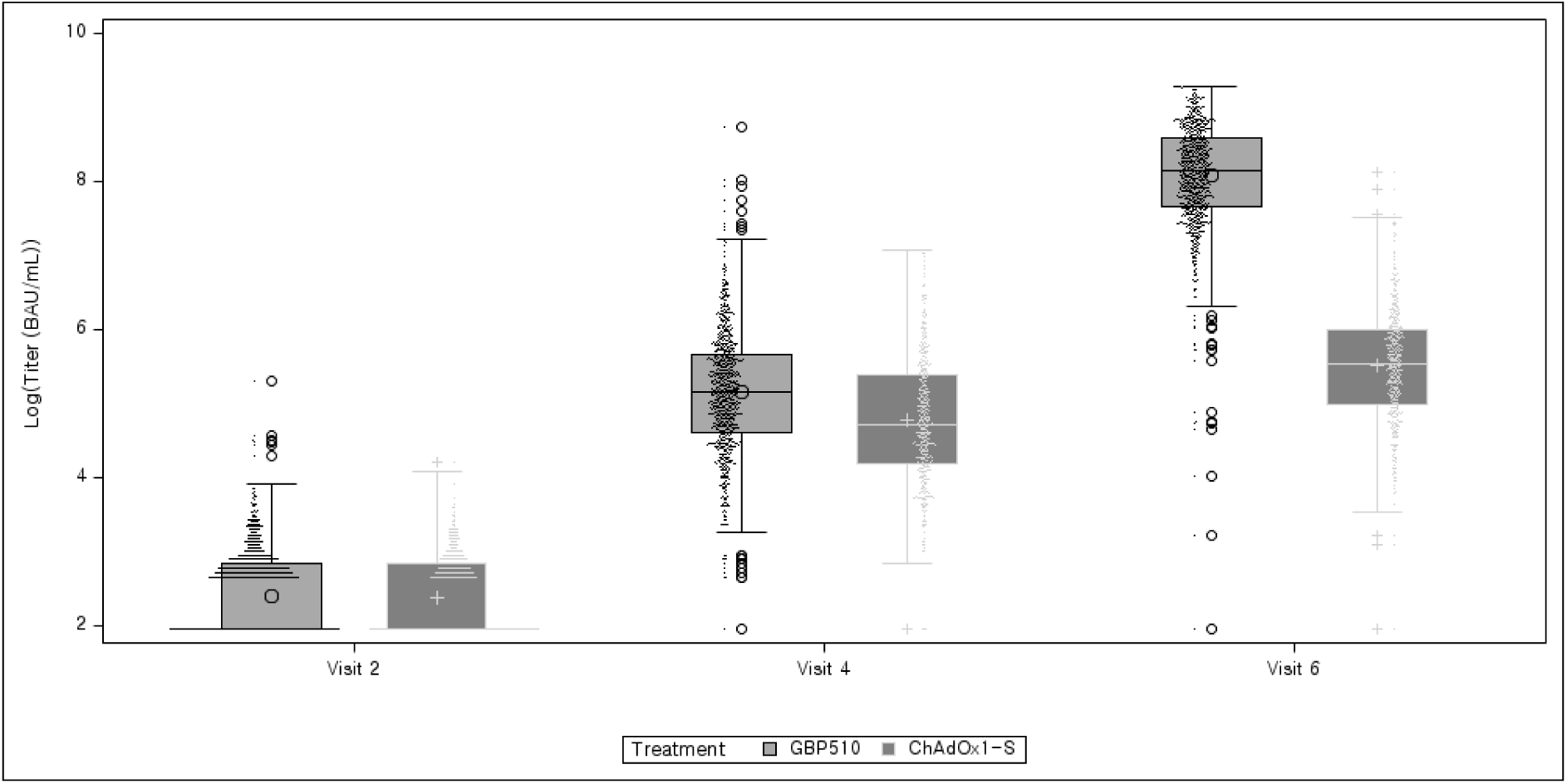
Boxplot for the Natural Logarithmic of Titre by ELISA at Visit 2, 4, and 6 (Per-Protocol Set 1)

**Figure S4.**
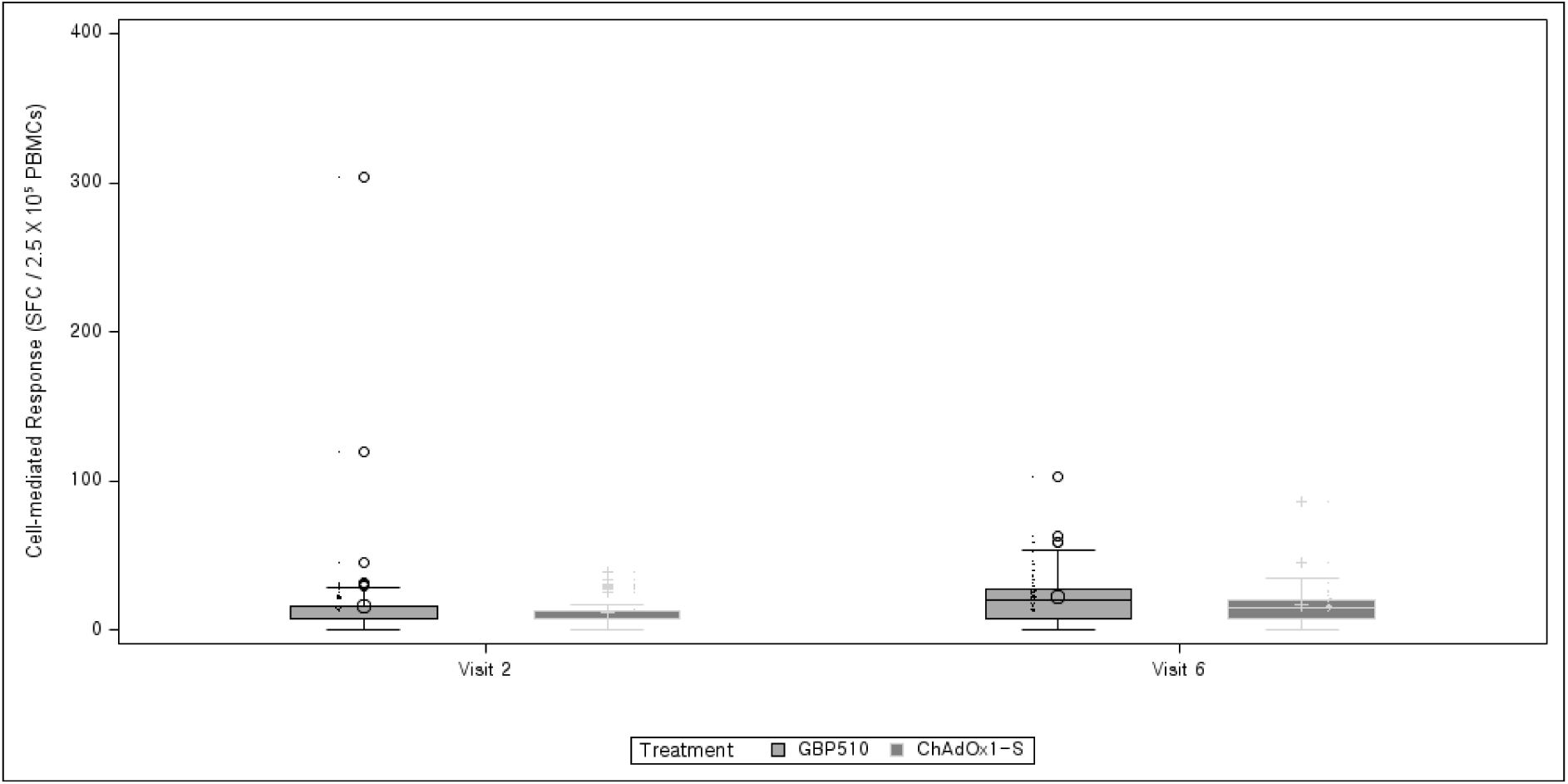
Cell-mediated Response FluoroSpot Assay Per-Protocol Set 1 [T cell: IFNγ]

**Figure S5.**
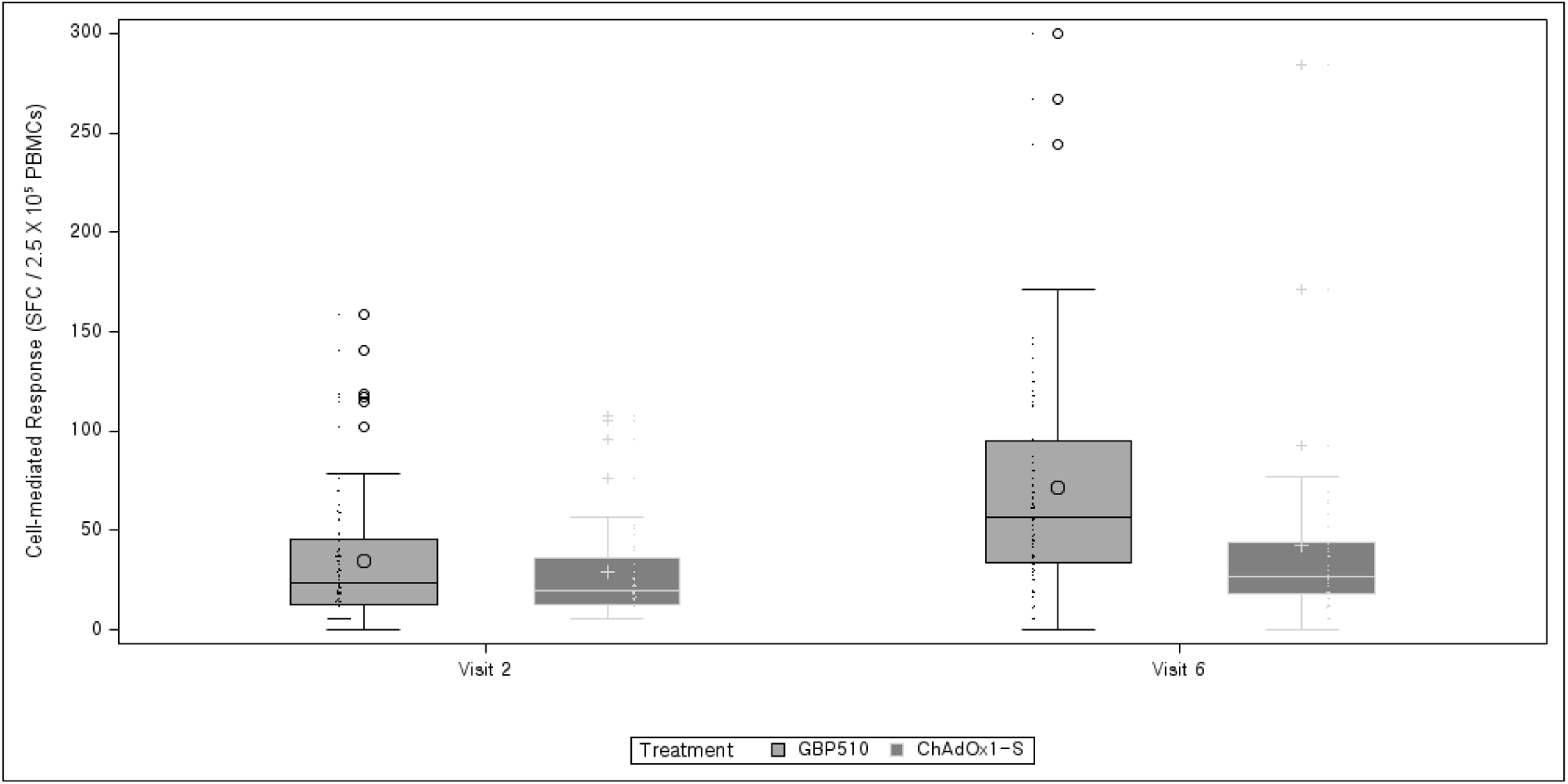
Cell-mediated Response FluoroSpot Assay Per-Protocol Set 1 [T cell: IL-2]

**Figure S6.**
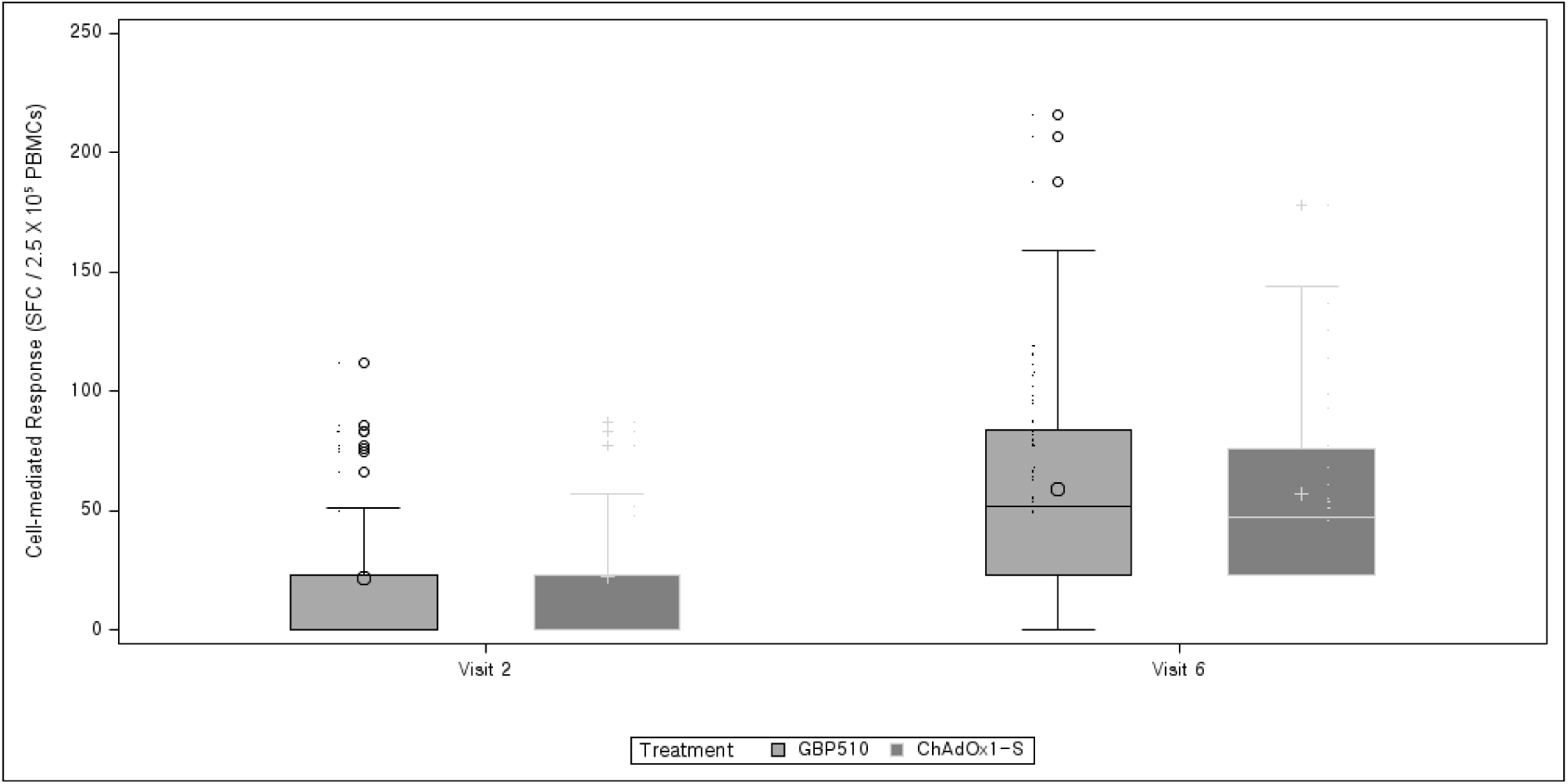
Cell-mediated Response FluoroSpot Assay Per-Protocol Set 1 [T cell: TNFα]

**Figure S7.**
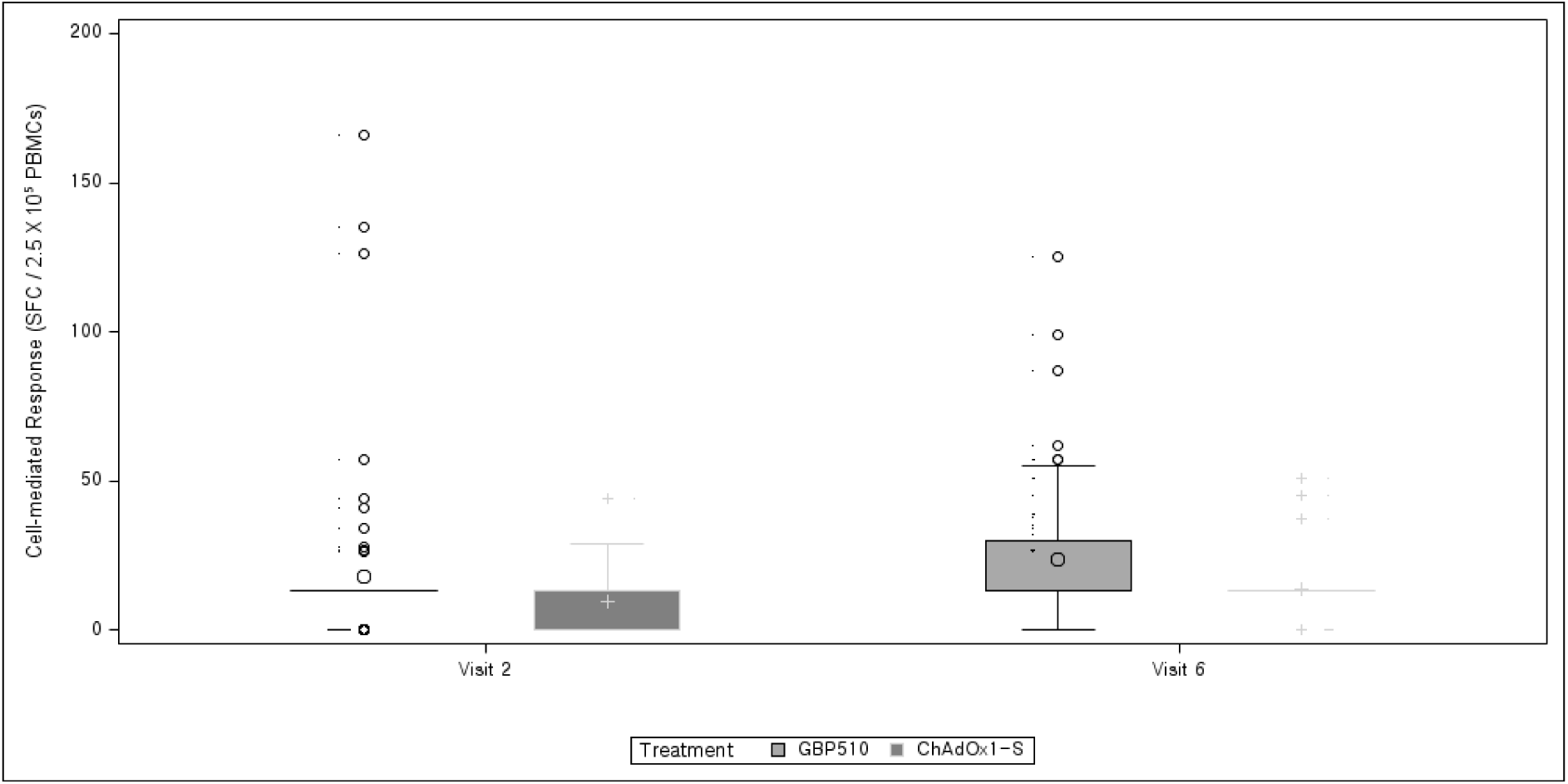
Cell-mediated Response FluoroSpot Assay Per-Protocol Set 1 [T cell: IL-4]

**Figure S8.**
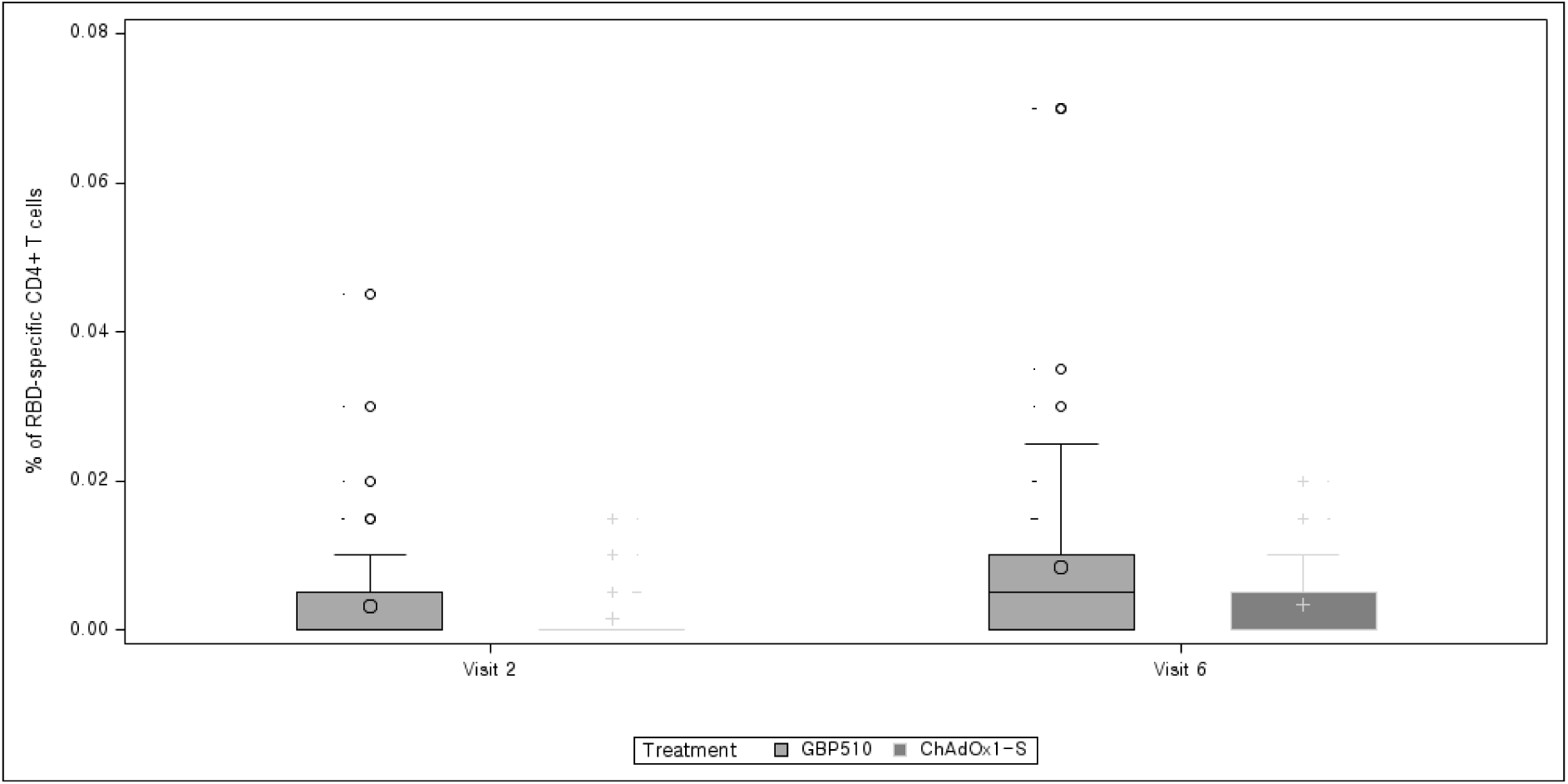
Cell-mediated Response FACS Per-Protocol Set 1 [CD4+ T cell: IFNγ]

**Figure S9.**
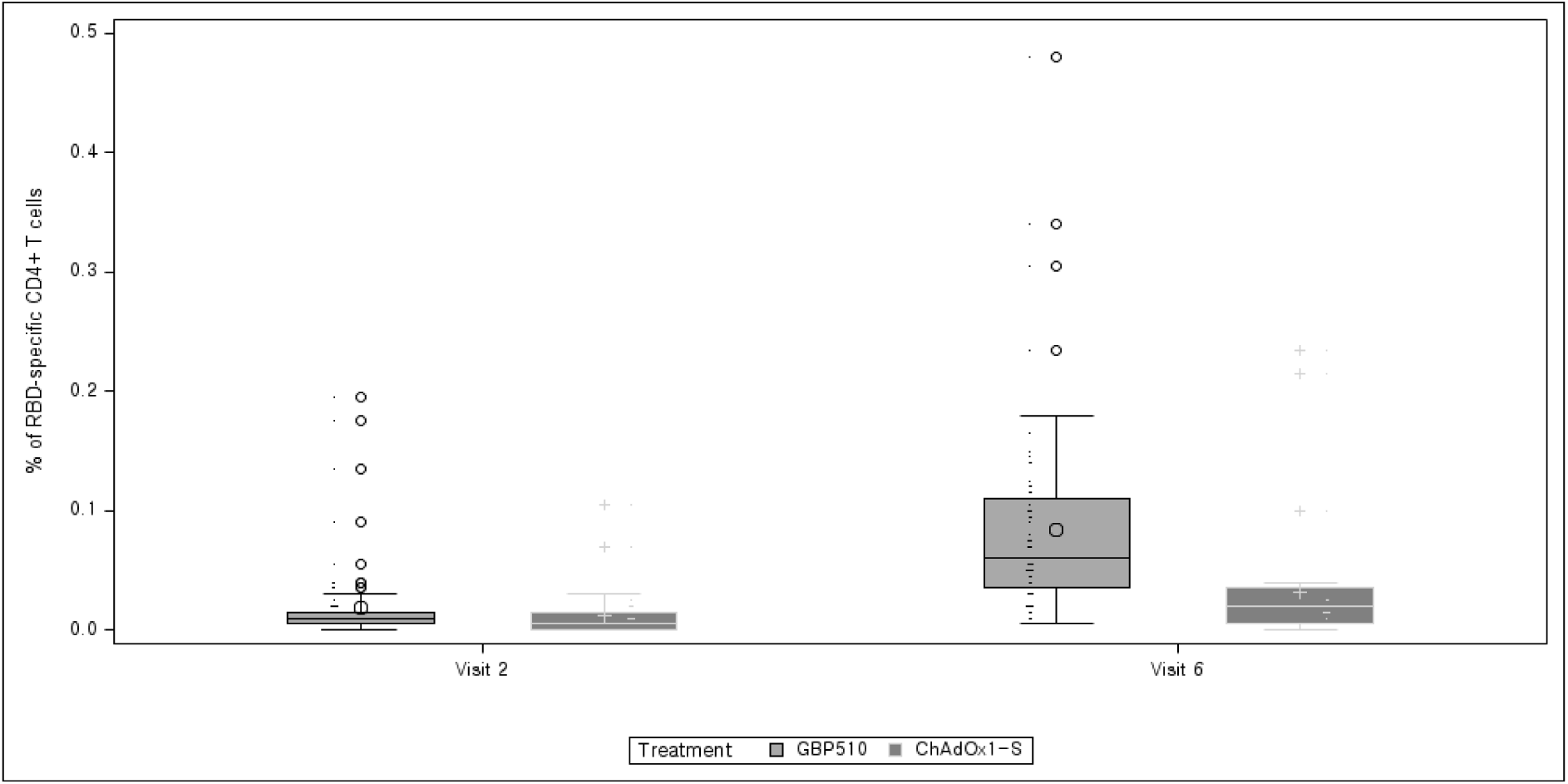
Cell-mediated Response FACS Per-Protocol Set 1 [CD4+ T cell: IL-2]

**Figure S10.**
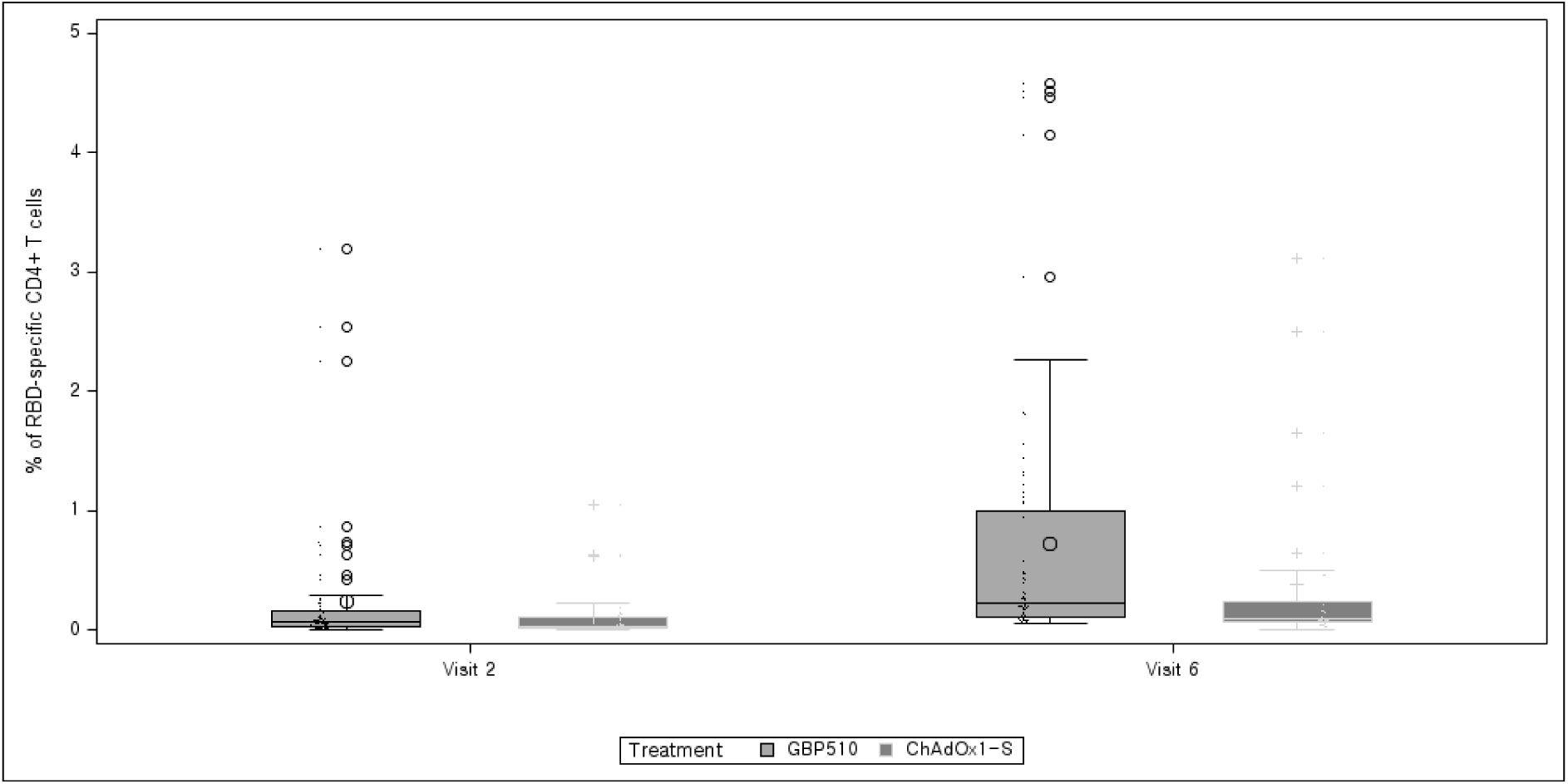
Cell-mediated Response FACS Per-Protocol Set 1 [CD4+ T cell: TNFα]

**Figure S11.**
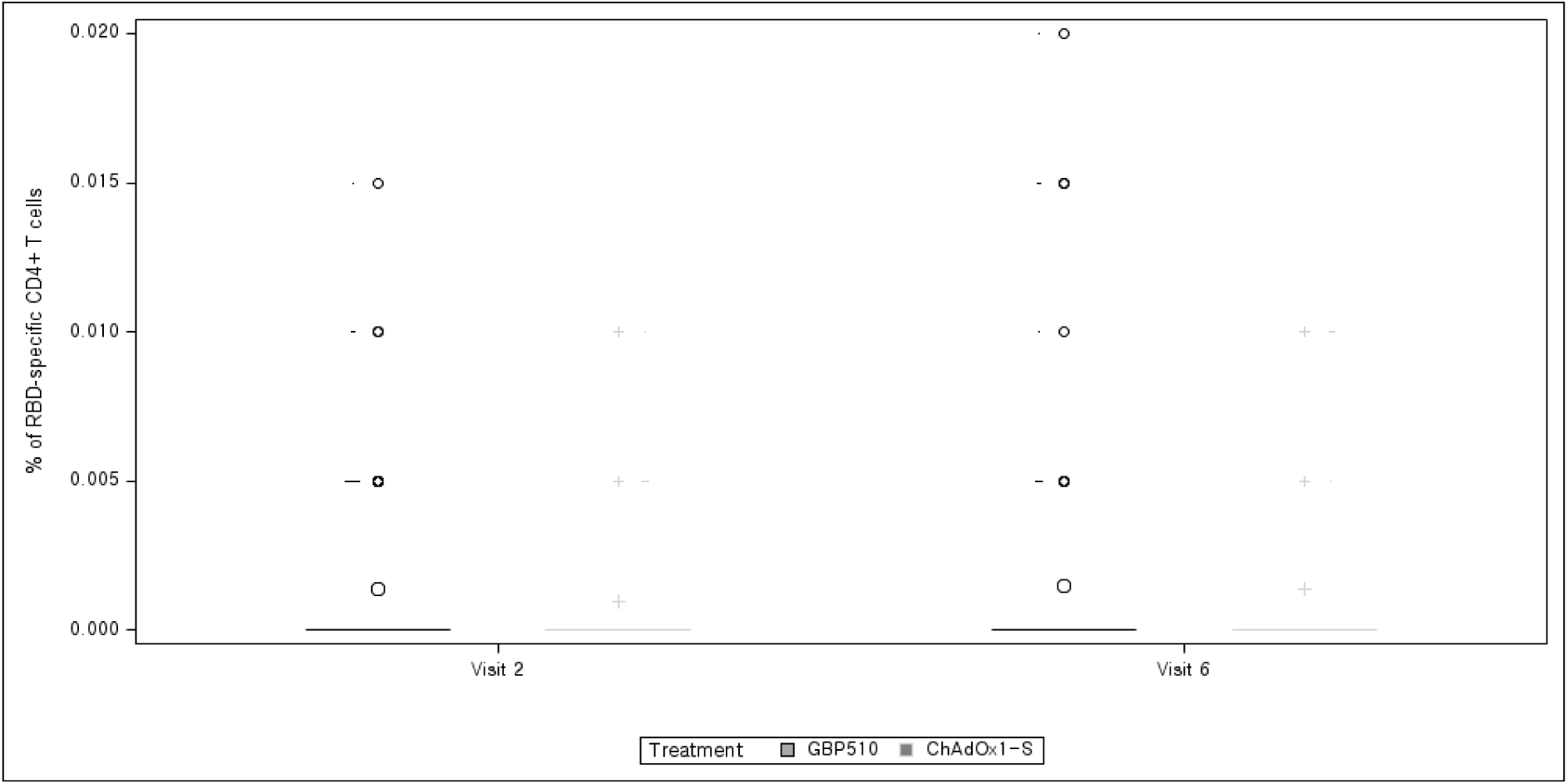
Cell-mediated Response FACS Per-Protocol Set 1 [CD4+ T cell: IL-4]

### III Supplementary Authors List

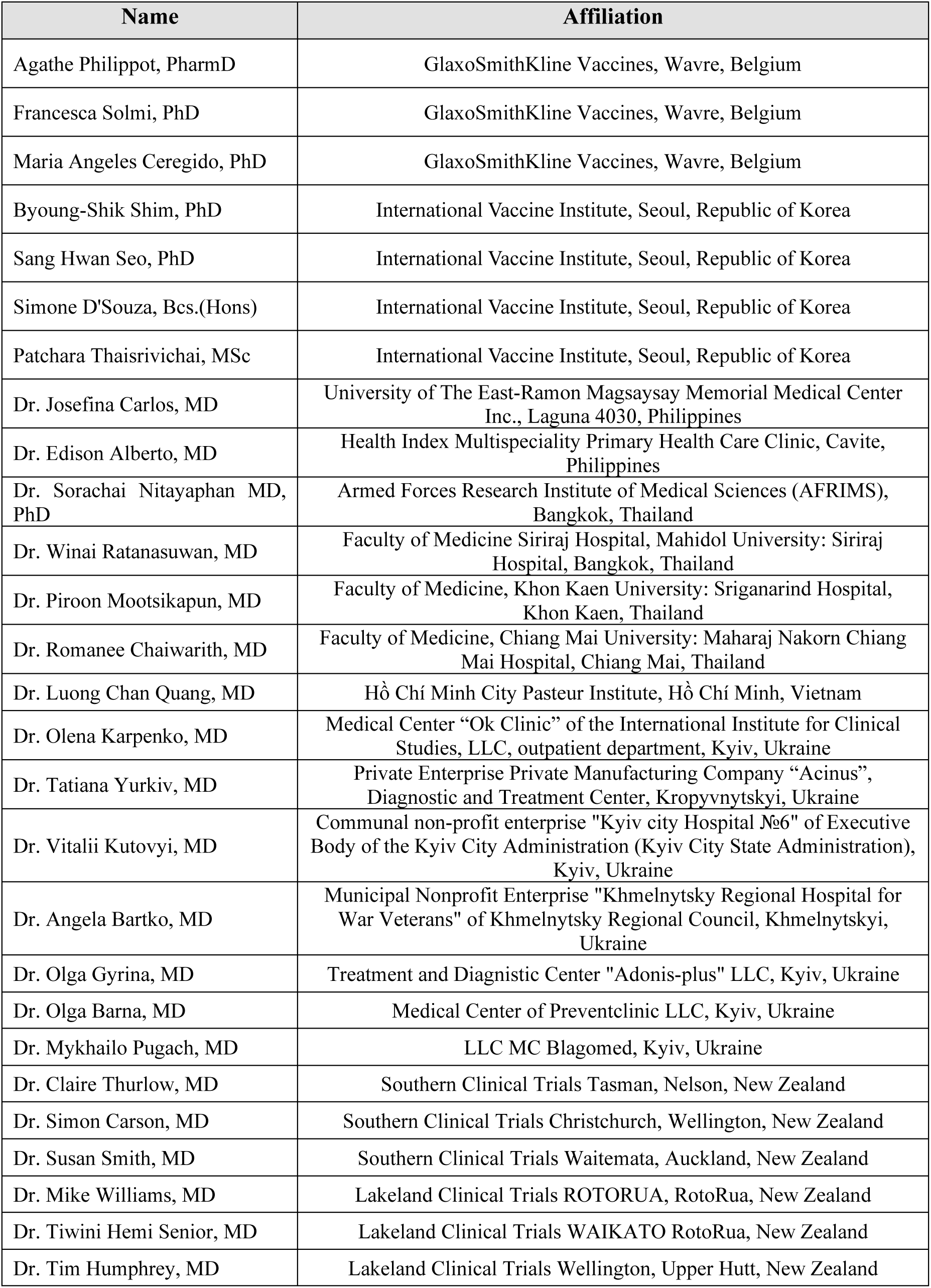
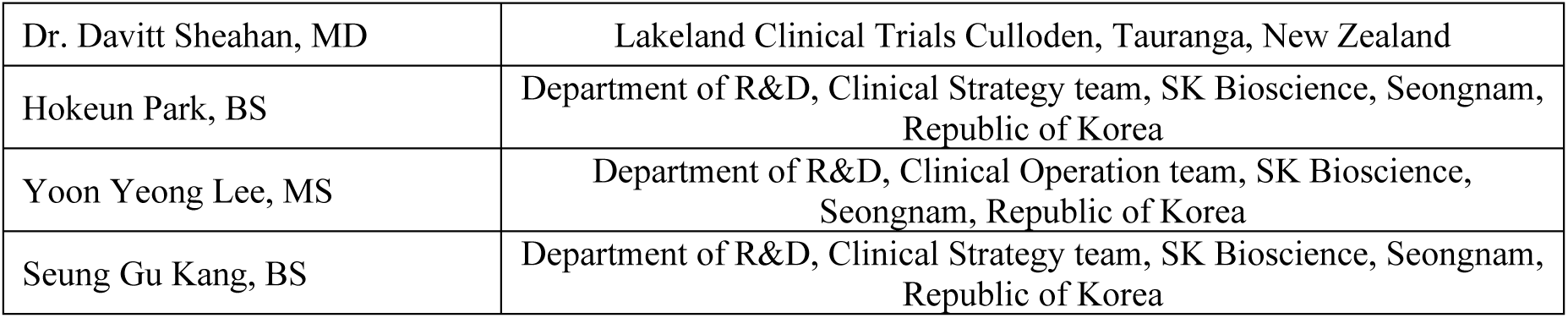

